# Bacterial co-infection and antimicrobial use in hospital-attended patients with laboratory-confirmed influenza infection: a systematic review and meta-analysis

**DOI:** 10.64898/2026.04.29.26352034

**Authors:** Harrison Bott, Ruonan Pei, Michael E Murphy, Ting Shi, Antonia Ho

## Abstract

**Background:** Bacterial co-infection contributes substantially to influenza-associated morbidity and mortality. Patterns of viral circulation, diagnostic testing and antimicrobial use changed markedly during the COVID-19 pandemic, yet contemporary estimates of bacterial co-infection and antimicrobial use in influenza have not been synthesised.

**Objectives:** To estimate the pooled prevalence of microbiologically confirmed bacterial co-infection among hospital-attended patients with laboratory-confirmed influenza. Secondary objectives were to characterise co-infecting bacterial pathogens, quantify antimicrobial prescribing overall and across key subgroups. This study was registered with PROSPERO (CRD420251072782).

**Data sources and eligibility:** We searched Embase (Ovid), MEDLINE, PubMed, Scopus, and Web of Science to 15th June 2025 for studies including ≥50 hospital-attended patients with laboratory-confirmed influenza and reporting bacterial co-infection.

**Methods:** Pooled prevalence estimates and antimicrobial prescription proportions were calculated using a generalised linear mixed model with logit link. Subgroup analyses included age group, clinical setting, and seasonal vs. pandemic influenza. Risk of bias was assessed using JBI Critical Appraisal tools and certainty of evidence using GRADE.

**Results:** Ninety-seven studies from 30 countries, comprising 116,273 patients with influenza, met inclusion criteria; 10,880 had confirmed bacterial co-infection. The pooled prevalence was 16.7% (95%CI 13.9-20.0%; I^2^=99.0%). Prevalence was higher in ICU compared to non-ICU settings (27.6% vs. 13.4%). The most frequently identified bacterial pathogens were *Streptococcus pneumoniae* (35.7%) and *Staphylococcus aureus* (30.3%). Antimicrobial use, reported in 38 studies, was high (pooled prevalence 88.1%, 95%CI 76.0-94.5%; I^2^=99.9%), and was more common in adults than children (97.8% vs 65.0%), and in ICU compared with non-ICU settings (96% vs 81%).

**Conclusions:** Bacterial co-infection was identified in approximately one in six hospital-attended influenza cases, yet antimicrobial prescribing is near-universal. Substantial heterogeneity and diagnostic variability constraint interpretation but underscore persistent challenges in clinical decision-making. These findings support strengthened diagnostic capacity and antimicrobial stewardship to optimise management of suspected influenza-associated bacterial co-infection.

## Introduction

Influenza remains a major global cause of respiratory illness, accounting for an estimated one billion infections and 290,000-650,000 respiratory deaths annually (1). The burden of disease is disproportionately focused on resource-limited settings, where 99% of influenza-related lower respiratory tract infection deaths, in children under five years, occur (2). Since 1900, four influenza pandemics have been documented, the most recent being the 2009 A(H1N1)pdm09 pandemic, which caused an estimated 201,200 respiratory deaths and 83,300 cardiovascular deaths worldwide (3). This pandemic disproportionately affected individuals under 65 years and 51% of these deaths occurred in Southeast Asia and Africa (4).

Bacterial co-infection has long been recognised as a major contributor of influenza-associated mortality. Analyses of the 1918 pandemic suggested that most deaths were attributable to secondary bacterial pneumonia (5). In contemporary clinical settings, bacterial co-infection is associated with greater disease severity, greater need for intensive care unit (ICU) admission, mechanical ventilation, and higher mortality (6–8).

However, diagnosing bacterial co-infection in patients with influenza remains challenging. Clinical features of viral and bacterial infection overlap, and existing microbiological testing may be limited by low sensitivity, delayed turnaround, or inconsistent use in routine practice (9). As a result, empirical antimicrobial therapy is frequently initiated in the absence of microbiological confirmation, with important implications for antimicrobial stewardship and the global spread of antimicrobial resistance (AMR) (10).

Previous systematic reviews have examined bacterial co-infection in influenza, but findings have been varied and often restricted to the 2009 pandemic period. Klein *et al.* reported a prevalence of up to 23% but did not report pooled estimates due to substantial heterogeneity (11), while Qiao *et al*. reported a pooled prevalence of 20.3% but included studies only up to 2021 (12). Notably, neither review systematically examined antimicrobial prescribing in relation to confirmed bacterial co-infection.

The COVID-19 pandemic significantly disrupted respiratory virus circulation, healthcare utilisation, diagnostic test practices, and antimicrobial prescribing behaviour (13–16). Updated evidence, including studies conducted during and after the pandemic, is therefore needed to understand bacterial co-infection dynamics in the context of these shifts, and to inform management strategies for future influenza pandemics. The 2024 update of the World Health Organization (WHO) Research Agenda for Influenza highlighted the identification of the frequency and aetiology of bacterial co-infection as a high priority in order to develop and implement optimal treatment strategies (17).

Accordingly, this systematic review and meta-analysis aimed to i) estimate the pooled prevalence of microbiologically confirmed bacterial co-infection among hospital-attended patients with laboratory-confirmed influenza, and ii) characterise the causative bacterial pathogens.

## Methods

This systematic review and meta-analysis were conducted in accordance with the Preferred Reporting Items for Systematic Reviews and Meta-Analyses (PRISMA) 2020 guidelines (18). The protocol was registered with PROSPERO (CRD420251072782).

We searched Embase (Ovid), MEDLINE, PubMed, Scopus, and Web of Science (including Science Citation Index Expanded, Conference Proceedings Citation Index, and Emerging Sources Citation Index), from database inception to 15th June 2025, without date restrictions. Studies that included at least 50 hospital attended patients with laboratory-confirmed influenza were eligible for inclusion. Full search strategies are provided in **(Supplementary Table 1).**

This review included studies that enrolled patients with laboratory-confirmed influenza and reported microbiologically confirmed bacterial co-infection within the study cohort. Patients without bacterial co-infection were not excluded from eligible cohorts, as their inclusion was necessary to estimate the prevalence of bacterial co-infection among individuals with influenza.□

Bacterial co-infection was defined as a microbiologically confirmed bacterial infection occurring in a patient with laboratory-confirmed influenza infection.

Studies reporting bacterial infection present at admission and/or secondary (hospital-acquired) bacterial infections developed during hospitalisation were eligible, provided all other eligibility criteria were met.

The widespread inconsistent reporting on the timing of microbiological testing and confirmation led us to include studies with confirmed bacterial co-infection of differing reported timepoints, and none.

Where it was reported, data on the testing methods and the corresponding sample sites were collected. Methods for bacterial identification included bacterial culture, Urinary Antigen Testing (UAT), PCR, and unspecified methods. Sample type or location include sputum, bronchoalveolar lavage (BAL), tracheal, pleural, bronchial, nasopharyngeal swab/aspirate, serum/blood, urine, and unspecified sites.

Similarly, for influenza testing, the methods included PCR, antigen testing, viral culture and unspecified methods. The sample sites included nasopharyngeal swab/aspirate, BAL, tracheal, bronchial, sputum, serum/blood and unspecified sites. A breakdown of testing can be found in Supplementary Tables 2-5.

Only studies conducted in hospital settings, including emergency departments, inpatient wards, and intensive care units (ICUs) were eligible for inclusion.

Two reviewers (HB and RP) independently screened titles, abstracts and full-text articles for eligibility. Discrepancies were resolved by discussion, or where necessary, consultation with a third reviewer (AH). Data were extracted using Covidence and independently verified by a second reviewer.

Extracted data included study characteristics (title, author, year of publication, journal, setting, study design, country or territory, and sample size), participant characteristics and diagnostic methods for influenza and bacterial infections. Where reported, influenza type and subtype were recorded. Bacterial pathogens were extracted at the species level, together with the proportion of patients with microbiologically confirmed bacterial co-infection. Data on antimicrobial therapy, including the proportion of patients receiving antibiotics, were also collected.

Risk of bias was independently assessed by two reviewers using the Joanna Briggs Institute (JBI) critical appraisal tools, with the tool selected according to study design (19). Disagreements were resolved by consensus. Risk of bias arising from missing results was not formally assessed.

The certainty of evidence for each outcome was evaluated independently by two reviewers using the “Grading of Recommendations Assessment, Development and Evaluation” (GRADE) approach (19). As detailed in the GRADE assessments (Supplementary Table 6a & 6b), publication bias was not formally assessed. Conventional methods, including funnel plots and Egger’s regression test, have limited utility in prevalence meta-analyses because substantial clinical and methodological heterogeneity can cause funnel plot asymmetry independent of selective publication. Given the extreme heterogeneity observed across analyses (I^2^= 99.0%), formal assessment of small-study effects was unlikely to provide interpretable or reliable evidence of publication bias.

The primary outcome was the pooled prevalence of microbiologically confirmed bacterial co-infection among hospital-attended patients with laboratory-confirmed influenza infection. Secondary outcomes included i) distribution of bacterial species; ii) prevalence of antimicrobial prescribing. Outcomes were stratified by age category, ICU vs. non-ICU setting, and pandemic vs. seasonal influenza.

The “pandemic influenza” period was defined as 2009-2017, corresponding to the years in which the WHO vaccine recommendations included A/California/7/2009 (20). Studies were classified as pandemic period if ≥50% of the study period fell within this interval. Using the same threshold, studies conducted predominantly during 2020-2023 were classified as during the COVID-19 pandemic period.

Pooled prevalence estimates of bacterial co-infection and antimicrobial prescribing were calculated using a generalised linear mixed model (GLMM) with logit link, generating 95% confidence intervals. Statistical heterogeneity was assessed using the I² statistic. Prespecified subgroup analyses were performed by age category, clinical setting (ICU vs. non-ICU), and pandemic vs. seasonal influenza. Sensitivity analyses were performed to assess the robustness of pooled estimates, including stratification by study design (prospective vs. retrospective). Potential sources of heterogeneity were explored using Cook’s distance, influence diagnostics and leave-one-out analyses.

All analyses were conducted using the *metafor* package in R version 4.5.3.

## Results

### Study selection and characteristics

A total of 1638 records were identified through database searches and a further 44 through citation searching. After removal of duplicates, 996 unique records underwent title and abstract screening, of which 97 studies met the inclusion criteria **(Figure 1)**.

**Figure 1.**
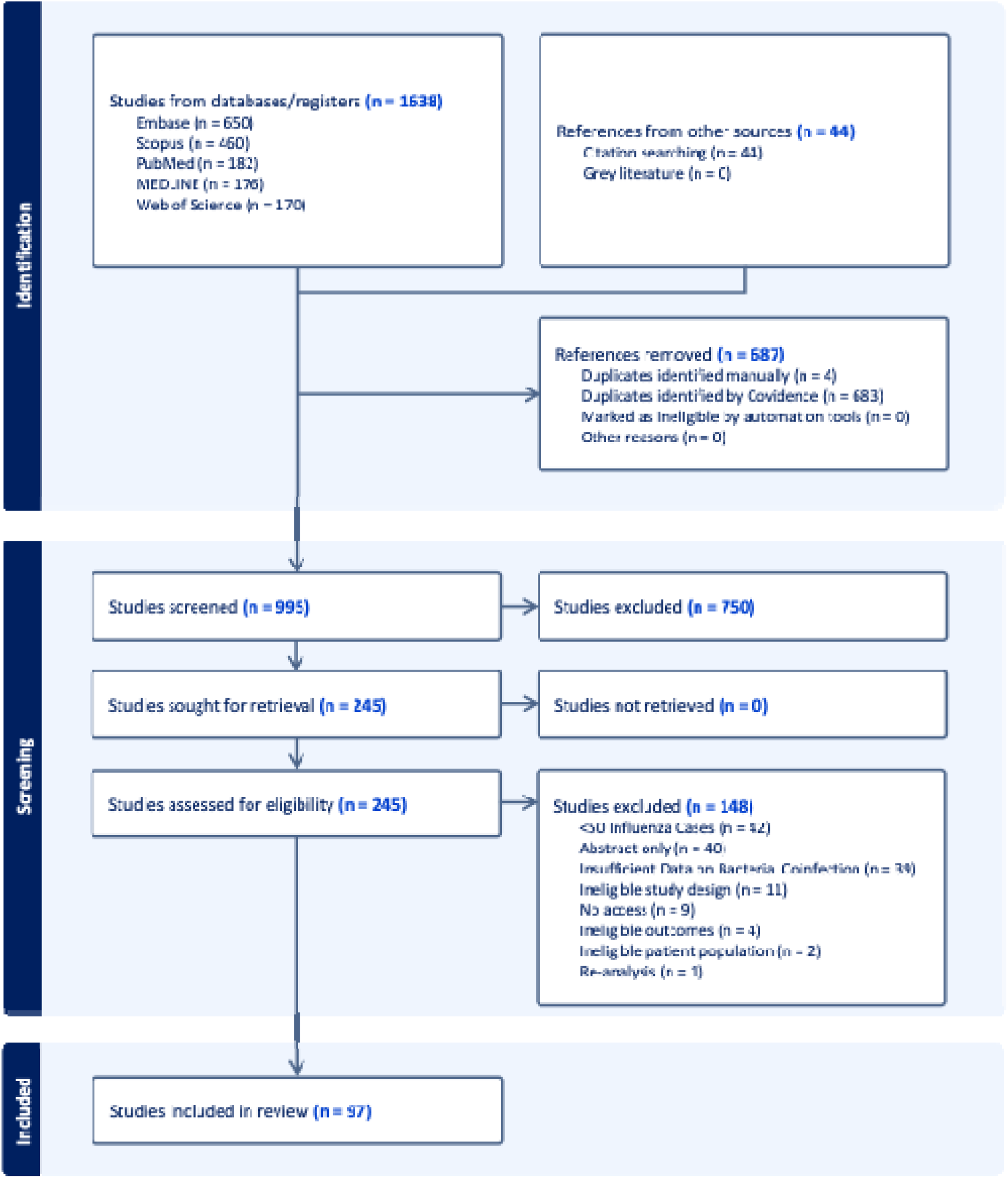
PRISMA flow diagram of study selection.

The included studies comprised 116,273 influenza patients from 30 countries and were published between 2009 and 2025 **(Figure 2)**. The largest number of studies were from the United States (n=15), China (n=15) and Spain (n=15). Most studies employed a retrospective design (n=62, 63.9%), while 35 (36.1%) were prospective. Fifty-six studies focused exclusively on adults, 29 in children, and 12 included participants of all age groups.

**Figure 2.**
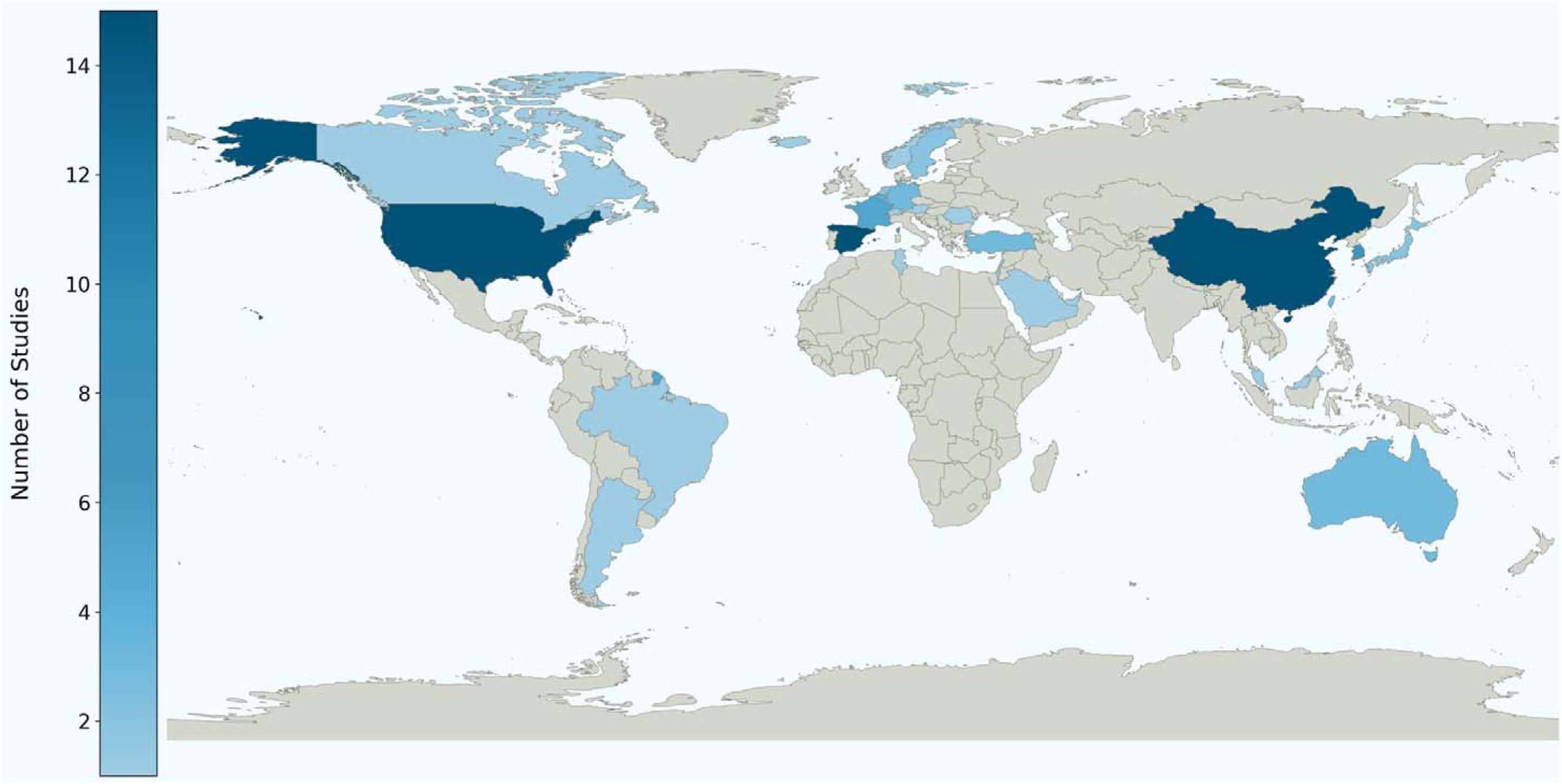
Visual representation of the geographic distribution of included studies by country/territory. Darker colour indicates higher number of studies. By WHO Region the geographic distribution of studies was African Region = 1, Eastern Mediterranean = 6, European Region = 40, Region of the Americas = 18, South-East Asian Region = 0, Western Pacific = 32.

Study size varied considerably. 25 studies enrolled 50-100 patients with influenza, 36 enrolled 101-500 patients, 16 enrolled 501-1000, 10 enrolled 1000-2500 patients, and 10 enrolled more than 2500 patients.

Thirty (30.9%) were conducted in ICU settings, while the remainder (n=67) were undertaken in emergency departments, hospital wards or mixed hospital settings **(Supplementary Table 7)**. Nearly three-quarters of included studies were conducted during the 2009 influenza A(H1N1) pdm09 pandemic period (71.1%, n=69) **(Supplementary Figure 2)**.

Influenza infection was most commonly confirmed by polymerase chain reaction (PCR) (n=83; 85.6%), 22 (22.7%) studies also reported using antigen testing, 12 (12.4%) reported using viral culture, while 6 did not clearly specify their Influenza testing methods (Supplementary Table 4).

Most studies (n=58, 59.8%) used samples obtained from the nasopharynx to identify influenza infection, including swabs and aspirate. Thirty-one (32.0%) did not specify sample location. **(Supplementary Table 5)**

Bacterial co-infection was predominantly identified by culture-based methods (n=72; 74.2%), with remaining studies used urinary antigen testing (UAT) (n=26,26.8%), PCR (n=15,15.5%), or did not specify the microbiological methods employed (n=19, 19.6%) **(Supplementary Table 2)**.

Information on microbiological sampling sites was reported in 79 studies (81.4%). Of these, 62 (78.5%) included lower respiratory tract (LRT) specimens, such as sputum, bronchoalveolar lavage (BAL), tracheal, bronchial or pleural samples). Sixteen studies (20.3%) used upper respiratory tract (URT) specimens, most commonly nasopharyngeal (NP) swabs, and 60 studies (76.0%) performed blood cultures. Two studies (2.5%) relied exclusively on URT samples for bacterial identification. The use of NP swabs was more common in paediatric cohorts (9/16 studies), reflecting the practical challenges associated with obtaining LRT samples from children. Consistent with this, sputum sampling was reported less frequently in paediatric studies (n=8) than in adult studies (n=32) **(Supplementary Table 3)**. Twenty studies included unspecified specimen types because sampling sites were not reported or incompletely described.

### Prevalence of bacterial co-infections

Reported prevalence estimates varied substantially across studies, ranging from 1.3% to 72.7%, reflecting heterogeneity in study populations, diagnostic approaches, and sampling practices. The lowest prevalence (1.3%) was reported by Hayashi *et al*. (21) in a retrospective analysis of 4491 non-ICU patients with influenza in Queensland, Australia. In contrast, Marcoux *et al.* (22) reported 72.7% co-infection prevalence among 55 influenza-positive patients in a retrospective, single-centre study in Belgium. This marked higher estimate is likely attributable to selection bias as the cohort consisted exclusively of ICU patients enrolled over an extended period to act as matched comparators for critically ill COVID-19 patients. Furthermore, these patients underwent extensive microbiological investigations for viral, bacterial and fungal pathogens, increasing likelihood of co-infection detection (22).

Across all included studies, the pooled prevalence of bacterial co-infection was 16.7% (95%CI 13.9-20.0%; I^2^=99.0%). **(Figure 3).** Age-stratified analysis showed no significant difference (p=0.238) between adult (n=56 studies; pooled prevalence 17.5%, 95%CI 14.1%-21.5%; I^2^= 98.7%) and children (n=29 studies; pooled prevalence 13.3%, 95%CI 8.7%-19.8%; I^2^=99.2%). Notably, a higher proportion of adult studies were conducted in ICU compared with paediatric studies (n=20; 66.7% vs. n=10; 33.3%). When analysis was restricted to ICU studies, the pooled prevalence was 26.6% (95%CI 21.0%-33.0%; I²= 97.3%) in adults and 32.4% (95%CI 21.8%-45.1%; I² = 92.0%) in children, with no significant difference (p=0.379) between age groups. Similarly, in non-ICU studies, pooled prevalence estimates were 13.7% (95%CI 10.3%-17.9%; I² = 98.6% in adults and 9.8% (95%CI: 6.0%-15.4%; I²= 99.2% in children, with no significant (p=0.226) between-group difference.

**Figure 3.**
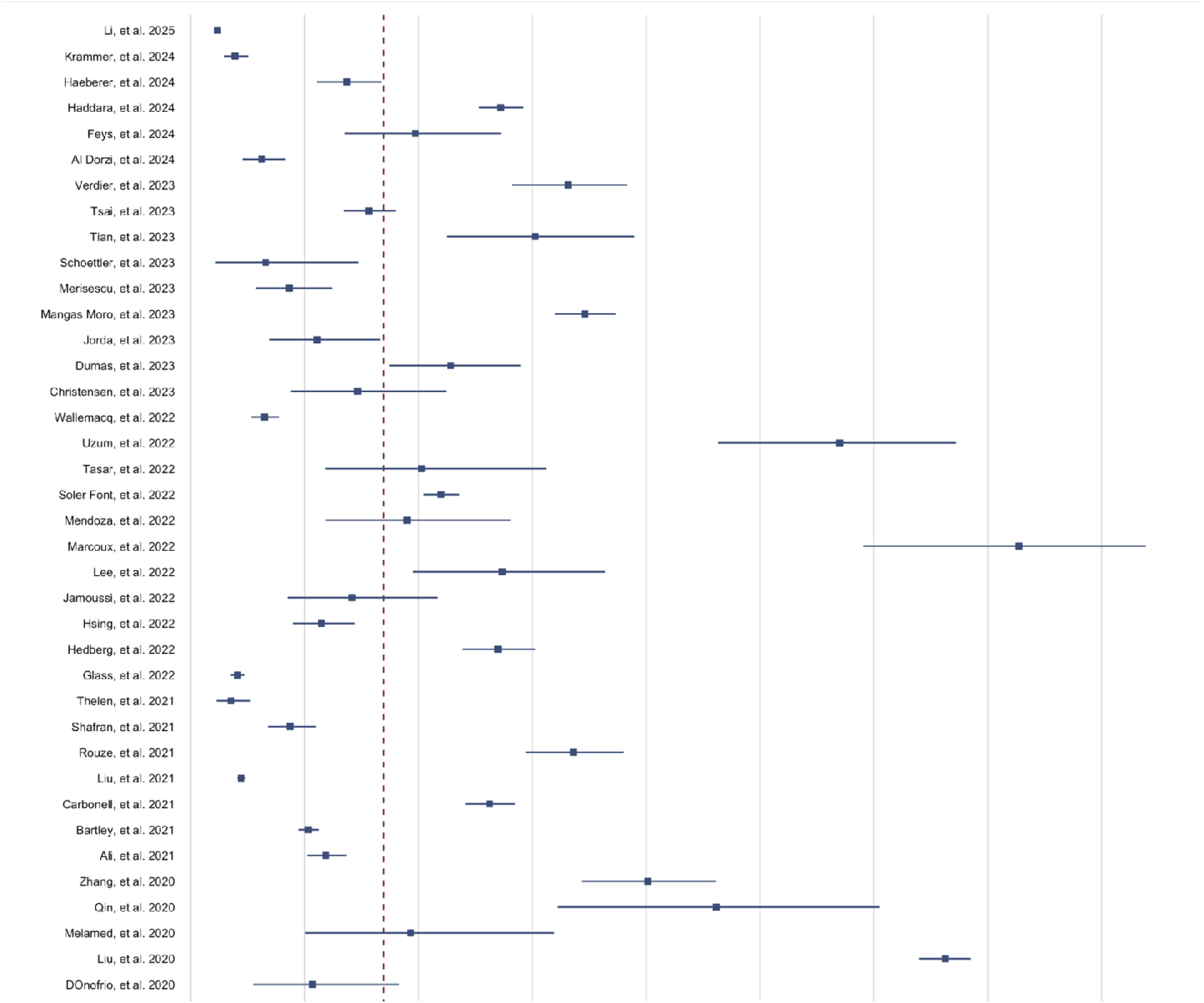
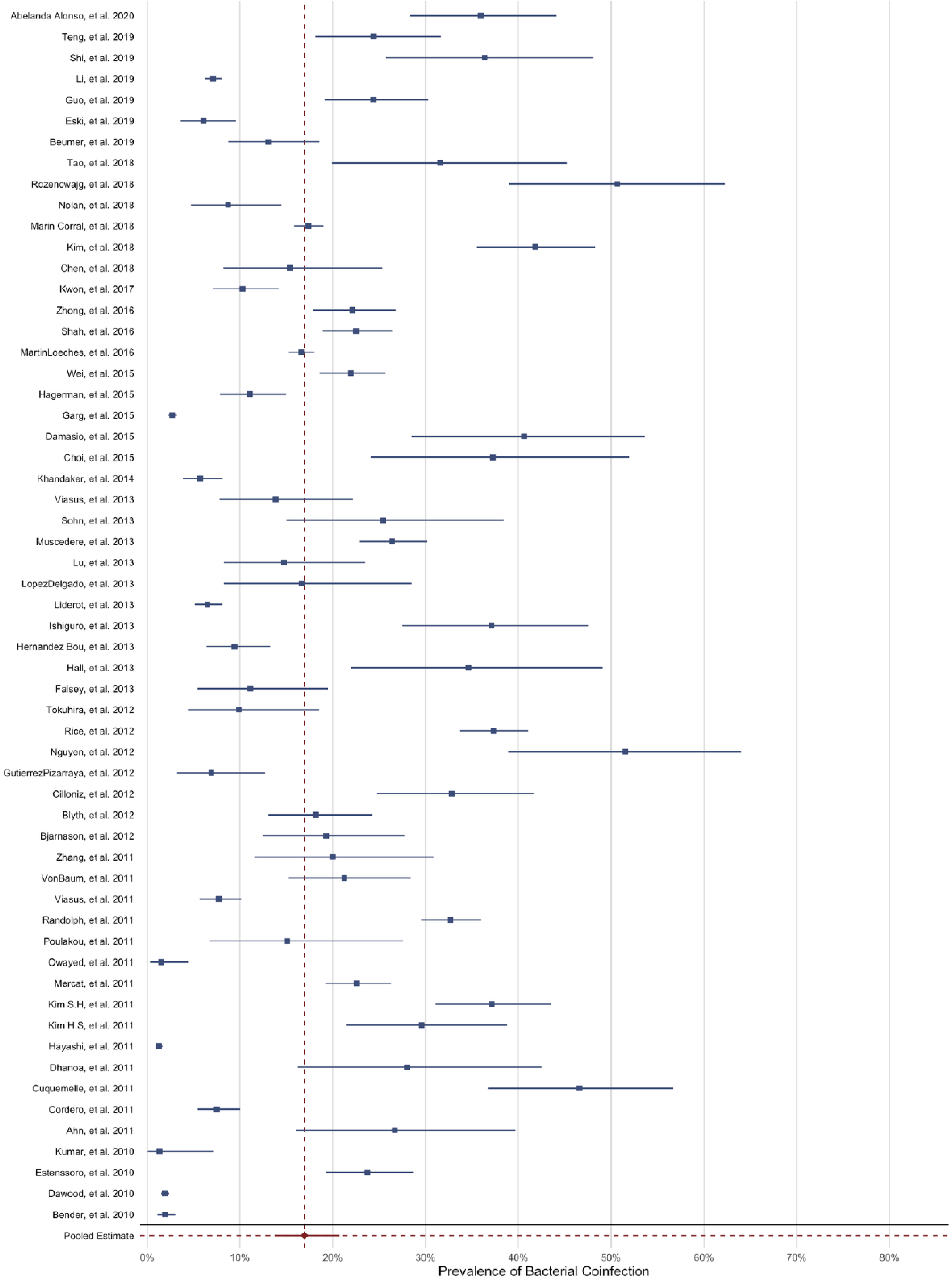
Forest plot of the prevalence of bacterial co-infection among patients with influenza. Each square represents the point estimate of bacterial co-infection prevalence from an individual study, with square size proportional to study weight in the meta-analysis. Horizontal lines indicate 95% confidence intervals (CIs). The pooled prevalence is shown by the diamond at the bottom of the plot. The dashed red line represents the 95% prediction interval, reflecting between-study heterogeneity and the expected range of prevalence in comparable settings.

Subgroup analyses demonstrated highly significant differences between pooled prevalence estimates in ICU settings (27.6%m 95%CI 22.9%-32.9%; I²= 97%) compared with non-ICU settings (13.2% (95%CI 10.4%-16.5%; I² = 99.0%; p <0.0001) **(Table 1)**.

**Table 1.** Pooled Prevalence of bacterial co-infection in hospital-attended influenza by sub-categories.

| Categories | N studies | Pooled Prevalence | 95%CI | P value | I² |
| --- | --- | --- | --- | --- | --- |
| All | 97 | 16.7% | 13.9-20.0% |  | 99.0% |
| Age Category |  |  |  |  |  |
| Adults | 56 | 17.5% | 14.1-21.5% | ns | 98.7% |
| Paediatrics | 29 | 13.3% | 8.7-19.8% |  | 99.2% |
| All ages | 12 | 22.4% | 14.0-32.6% |  | 98.0% |
| Clinical Setting |  |  |  |  |  |
| ICU | 30 | 27.6% | 22.9-32.9% | <0.0001 | 97.0% |
| Non-ICU | 67 | 13.2% | 10.4-16.5% |  | 99.0% |
| Pandemic Status |  |  |  |  |  |
| Seasonal | 28 | 13.3% | 8.9%-19.6% | ns | 99.3% |
| Pandemic | 69 | 18.4% | 15.1%-22.2% |  | 98.6% |

When stratified by influenza period, the pooled prevalence was 13.3% (95%CI 8.9%-19.6%; I²= 99.3% for seasonal influenza and 18.4% (95%CI 15.1%-22.2%; I²=98.6%) for pandemic influenza, with no significant difference (p=0.142) between groups.

In a sensitivity analysis, studies were stratified by design as prospective or retrospective. The pooled prevalence of bacterial co-infection was 19.3% in prospective studies (95%CI 15.6%-23.5%; I²=96.3%) and 15.5% among retrospective studies (95%CI 11.9%-19.9%; I²=99.3%), with no significant difference by study design (p=0.190).

### Bacterial aetiology

Fifty-nine (60.8%) studies reported bacterial pathogens at the species level. Overall, *Streptococcus pneumoniae* (n=1191; 35.7%) and *Staphylococcus aureus* (n=1012; 30.3%) predominated accounting for approximately two-thirds of identified co-infections, with *Haemophilus influenzae* (n=275; 8.3%) and *Pseudomonas aeruginosa* (n=227; 6.8%) each contributing less than 10% **(Figure 4).**

**Figure 4.**
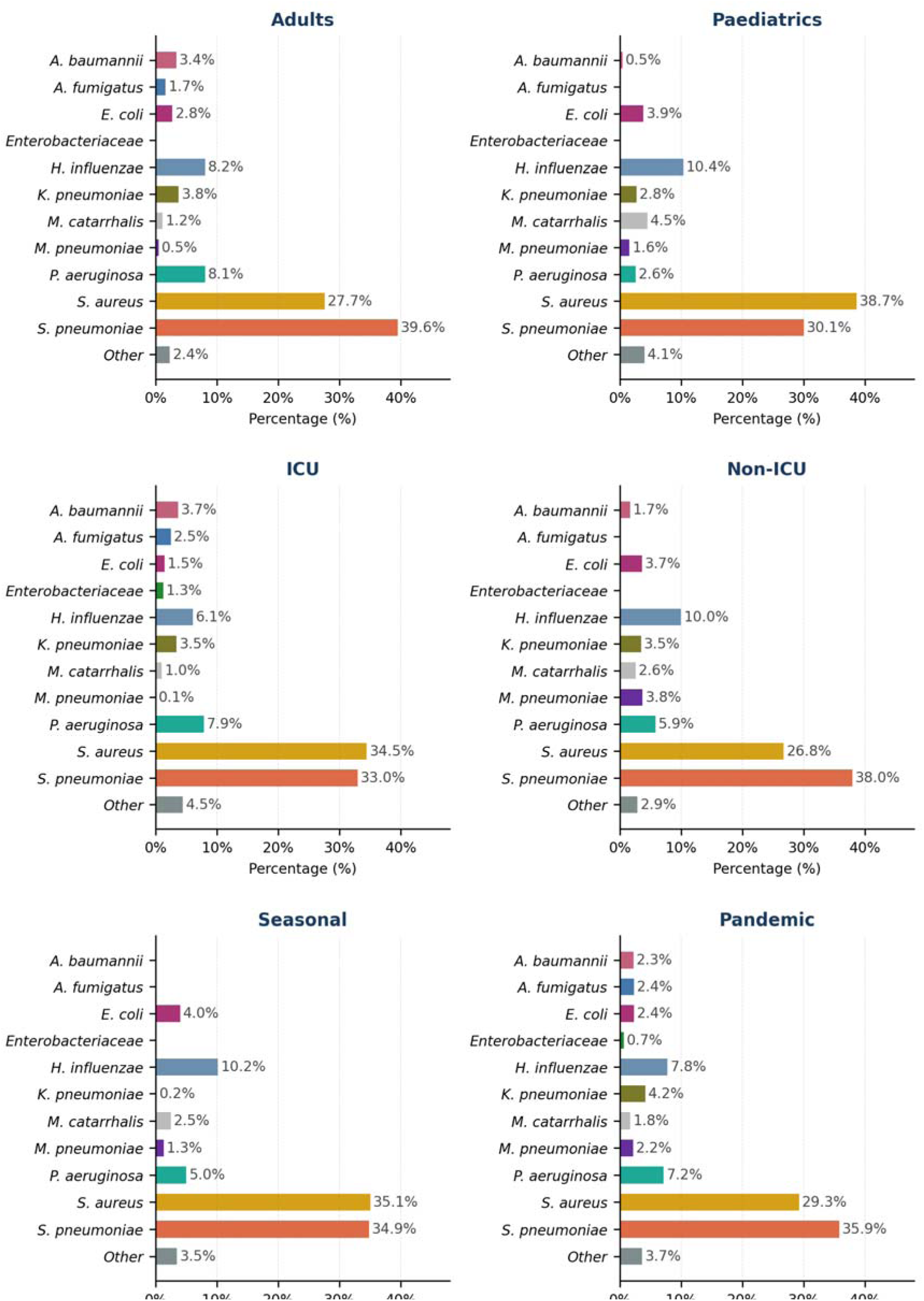
Distribution of bacterial pathogens among patients with influenza stratified by population and setting. Bar charts depict the proportional distribution of bacterial pathogens identified in patients with hospital-attended influenza across predefined subgroups: Adults, Paediatrics, Intensive Care Unit (ICU), Non-ICU, Pandemic influenza, and Seasonal influenza. In Adults, non-ICU, and Pandemic the most frequently identified pathogen was S. pneumoniae, in Paediatrics, ICU, and Seasonal it was aureus. Percentages represent the relative frequency of each pathogen among reported bacterial co-infections within each subgroup.

Pathogen distributions varied modestly by age group, clinical setting, and influenza period. Adult-focused and non-ICU studies were most commonly dominated by *S. pneumoniae* (39.6% and 38.0%, respectively), *whereas S. aureus* were most frequently identified in paediatric population (38.7%), ICU settings (34.5%) and seasonal influenza (35.1%) studies.

Across all strata, the same small group of pathogens accounted for the majority of bacterial co-infections, with limited variation in relative proportions.

### Antimicrobial prescribing

Across all studies reporting antimicrobial usage (n=38), prescribing rates ranged from 4.7% to 100%, variability reflected differences in study populations and reporting practices **(Figure 5).** The lowest rate (4.7%) was reported by Liderot *et al*. in a Swedish tertiary hospital, where the prevalence of co-infection was 6.5% (23). Among adult-focused studies reporting antimicrobial use (n=18), six (33%) prescribed antibiotics to all patients (24–29). All but one study was from Europe and were evenly split between ICU and non-ICU settings.

**Figure 5.**
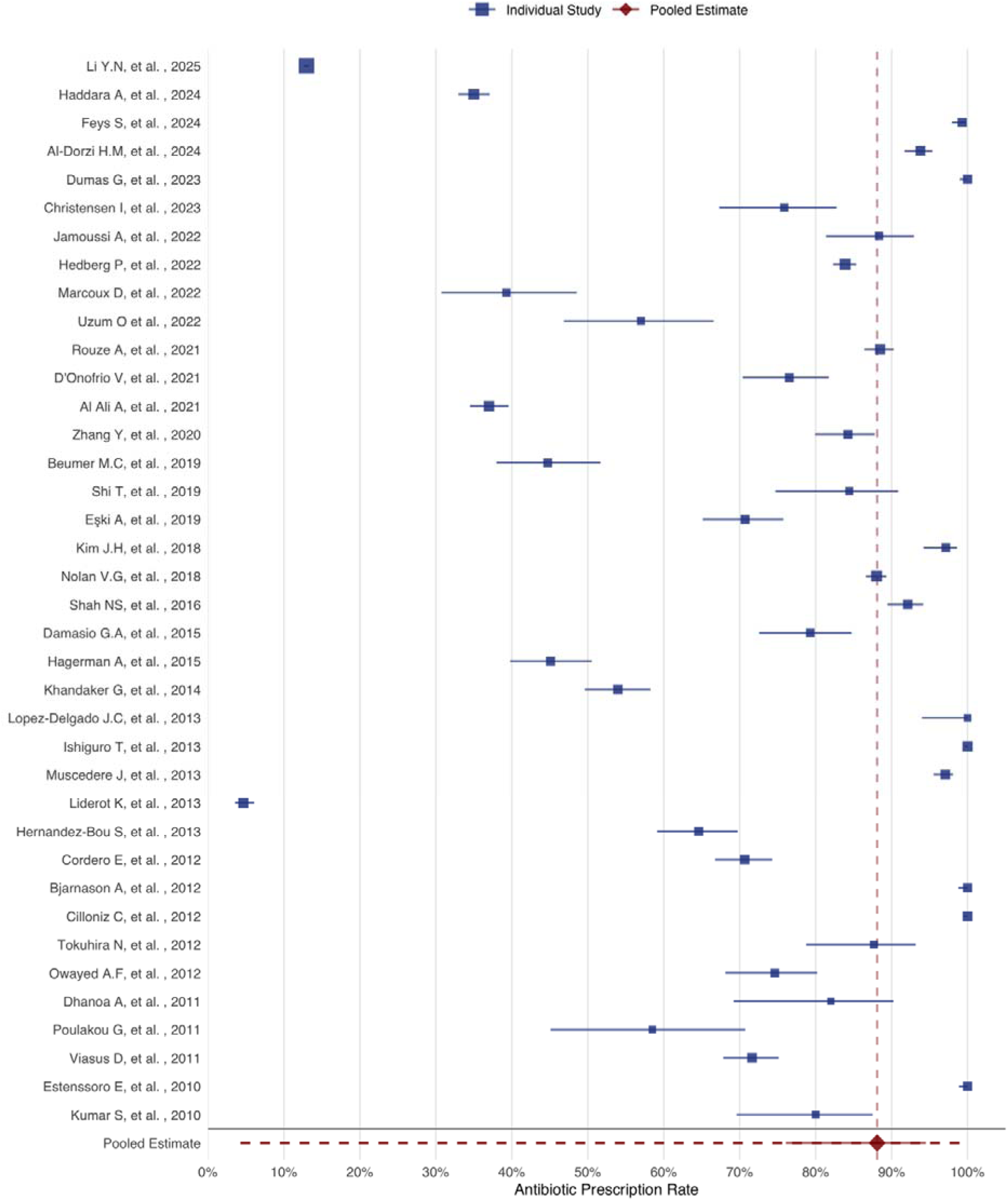
***Forest plot of the prevalence of antibiotic prescribing in included studies*** Squares represent the point prevalence of antimicrobial prescribing reported by individual studies, with horizontal lines indicating the corresponding 95%CIs. The red diamond represents the pooled prescription prevalence estimate is. The vertical dashed red line denotes the overall pooled proportion of antibiotic prescribing. Studies are presented chronologically by publication year along the y-axis, and the x-axis represents the proportion of patients receiving antimicrobials (%).

Overall, 23 studies prescribed antimicrobials to at least 75% of their patient cohorts. The pooled prevalence of antimicrobial prescription was 88.1% (95%CI, 76.1%-94.5%; I^2^=99.9%), substantially exceeding the prevalence of confirmed bacterial co-infection. Antibiotic prescription was higher in adults vs. children (97.8% vs 65.0%; p=0.0017) and in ICU, compared to non-ICU settings (96% vs 81%; p=0.049), but prevalence was not significantly different between pandemic and seasonal influenza studies **(Table 2)**.

**Table 2.** Pooled prevalence of antimicrobial prescription stratified by subgroup.

| Categories | N studies | Pooled Prevalence | 95%CI | P value | I<sup>2</sup> |
| --- | --- | --- | --- | --- | --- |
| All | 38 | 88.1% | 76.1-94.5% |  | 99.9% |
| Age Category |  |  |  |  |  |
| Adults | 18 | 97.8% | 87.0-99.7% | 0.0017 | 99.8% |
| Paediatrics | 12 | 64.7% | 49.4-77.4% |  | 99.5% |
| Clinical Setting |  |  |  |  |  |
| ICU | 12 | 96.2% | 84.8-99.1% | 0.049 | 99.0% |
| Non-ICU | 26 | 80.8% | 62.1-91.5% |  | 99.9% |
| Pandemic Status |  |  |  |  |  |
| Seasonal | 11 | 84.3% | 40.3-97.7% | ns | 99.9% |
| Pandemic | 27 | 89.3% | 78.4-95.0% |  | 99.6% |

### Studies undertaken during the COVID-19 pandemic

Four studies, including 44,145 patients, were conducted predominantly during the COVID-19 pandemic (30–33). All were conducted in non-ICU settings (two adults and two paediatric). Reported bacterial co-infection prevalence ranged from 2.3% to 8.6%, and antimicrobial prescribing ranged from 13.0% to 93.8%.

### Risk of bias and certainty of evidence

The JBI critical appraisal tools were used to assess the risk of bias in included studies, with the tool selected according to the design of each study **(Supplementary Table 8-12).** Consistent with JBI guidance, these tools do not generate an overall quality score or summary risk-of-bias classification; rather, they identify potential sources of bias across individual methodological domains.

Several recurring limitations were evident across the included studies. Most notably, many studies did not adequately identify or account for potential confounding factors. In addition, few studies established temporal precedence between viral infection and bacterial co-infection, making it difficult to determine whether bacterial pathogens were present at hospital admission or study enrolment, or was acquired subsequently. This uncertainty largely reflects heterogeneity in the timing and methods of microbiological testing across studies. These limitations are closely linked to the predominance of retrospective study design, which restricted the ability to determine the sequence of infection events and to address confounding through prospective data collection and analysis.

Using GRADE, the certainty of evidence was rated as very low for the primary outcome and for the proportion of patients with confirmed bacterial co-infection who received antimicrobials **(Supplementary Table 6a & 6b).** This was primarily attributable to serious risk of bias from non-systematic microbiological testing and clinician-driven empirical prescribing, extreme unexplained heterogeneity, serious indirectness related to diagnostic variability and timing of testing, incomplete microbiological evaluation, and inability to link antimicrobial use to microbiologically confirmed infection at the individual patient level.

Influence diagnostics identified five highly influential studies **(Supplementary Figure 1)**. However, leave-one-out and joint exclusion analyses resulted in minimal changes to pooled estimates and heterogeneity, indicating that no single study unduly influenced the overall findings.

Many included studies reported microbiologically confirmed bacterial co-infection but failed to specify their methods of identification and/or their sample site. These studies reported confirmed infections and were reviewed individually and assessed during quality appraisal to ensure they still met eligibility criteria.

## Discussion

This systematic review and meta-analysis provide the most up-to-date synthesis of evidence of bacterial co-infection in hospital-attended patients with influenza. Among more than 116,273 patients across 97 studies, bacterial co-infection was identified in approximately 16.7% of cases, although estimates varied considerably. Co-infection prevalence was consistently higher in ICU populations, reflecting greater illness severity, more intensive diagnostic investigation, and ICU-specific risks such as prolonged hospitalisation, invasive devices and mechanical ventilation. Consequently, ICU prevalence estimates likely represent a mixture of early community-acquired co-infections, later secondary or hospital-acquired infections, or ventilator-associated pneumonia (34), which could not be reliably disentangled in aggregate analyses due to limited reporting of sampling time.

The predominant bacterial pathogens, *S. pneumoniae, S. aureus, H. influenzae* and, *P. aeruginosa*, are consistent with established evidence linking these organisms to severe influenza-associated pneumonia (35–37). However, interpretation is complicated by heterogeneous sampling strategies and the potential for colonisation, particularly for *S. aureus* and Gram-negative organisms in ventilated patients. Many studies relied on blood culture, urinary antigen testing, did not specify sample type, or used URT samples, mainly in paediatric studies where lower respiratory sampling is challenging, further limiting inference as nasopharyngeal detection may represent colonisation rather than true lower respiratory tract infection (38, 39).

The primary objective of this review was to estimate the prevalence of bacterial co-infection among patients with influenza and to characterise the causative bacterial pathogens. Although these findings provide important epidemiological insights, they do not identify which patients are at highest risk of bacterial co-infection, which may be of greater clinical relevance for guiding antimicrobial prescribing. However, demographic, clinical, and laboratory predictors of bacterial co-infection were inconsistently reported across studies, precluding robust synthesis. Future prospective studies using standardised definitions and data collection are needed to support the development of risk-stratification approaches. Interpretation of co-infection prevalence is further complicated by real-world microbiological sampling practices. The timing of specimen collection relative to empirical antimicrobial initiation was rarely reported, despite its likely impact on diagnostic yield. In routine care, antibiotics are often initiated before microbiological samples are obtained, potentially suppressing bacterial growth and reducing culture positivity. Incomplete or absent microbiological sampling, especially among patients with less severe disease, may further contribute to under-ascertainment of bacterial co-infection. Although these practices reflect pragmatic clinical decision-making, they introduce uncertainty into prevalence estimates derived from observational studies.

The predominance of culture-based diagnostics, which are less sensitive than molecular approaches (40, 41), may also have contributed to the underestimation of true bacterial burden. Moreover, the absence of microbiological confirmation does not exclude bacterial infection, and clinicians frequently diagnose bacterial co-infection using a combination of clinical, radiological and laboratory findings. However, such approaches are heterogeneous and difficult to standardise across studies. We therefore restricted our analysis to microbiologically confirmed co-infection, a more objective and consistently measurable outcome. Consequently, the pooled estimates should be interpreted as estimates of microbiologically confirmed bacterial co-infection rather than the total burden of clinically diagnosed bacterial infection in patients with influenza. Although radiological findings remain central to the diagnosis of pneumonia, microbiological confirmation provides important information on pathogen epidemiology and AMR, with direct implications for treatment decisions and antimicrobial stewardship.

Despite the relatively modest prevalence of confirmed bacterial co-infection, antimicrobial prescribing was near universal, with antibiotics administered to 88.1% of patients. Interpretation of antimicrobial use is constrained by limited reporting, typically as a binary outcome without detail on indication, duration, or timing relative to influenza diagnosis and microbiological sampling. As a result, defining “appropriate” antimicrobial use using retrospective and aggregate data alone is not possible. High prescribing rates may overestimate inappropriate exposure if empirical therapy was discontinued following influenza confirmation, while the absence of patient-level linkage precludes assessment of whether antibiotics were appropriately targeted, de-escalated, or unnecessarily continued.

Our pooled prevalence estimates are marginally lower than those reported in earlier reviews (11, 12), likely reflecting methodological differences and inclusion of more recent studies up to 2025. Notably, high antimicrobial use persists despite increased availability of molecular diagnostics and greater awareness of AMR, mirroring patterns observed in both seasonal influenza and COVID-19. The substantially lower prevalence of bacterial co-infections reported in hospitalised COVID-19 patients (6.9%, 95%CI 4.3-9.5%) and differences in prominent pathogens (*Mycoplasma spp.*, *H. influenzae* and *P. aeruginosa* reported most frequently), underscore the importance of early, accurate viral diagnosis at admission to support more targeted antimicrobial prescribing (42).

Heterogeneity across analyses was very high, reflecting variation in study design, patient populations, illness severity, diagnostic methods, and sampling practices. Therefore, pooled prevalence estimates reported in this review should be interpreted as a descriptive synthesis across highly heterogeneous settings and clinical methodologies rather than as a universally applicable estimate. Geographical representation was also uneven, with sparse data from Africa and South-East Asia, limiting global generalisability.

Together, these findings highlight the challenge of balancing timely empirical therapy with antimicrobial stewardship in influenza. Improved reporting of sampling indications, timing relative to antimicrobial initiation, and treatment duration would substantially enhance interpretability. Future research should prioritise prospective studies with standardised, protocolised microbiological testing and detailed treatment timelines to better distinguish true bacterial infection from colonisation, secondary infection and empirical over-treatment. Expanding access to rapid, reliable point-of-care diagnostics, alongside strengthened stewardship and prescriber support, will be essential to optimise care for patients with influenza and reduce unnecessary antimicrobial exposure during both seasonal epidemics and future respiratory viral pandemics.

This review has several limitations that should be considered when interpreting its findings. First, the included studies exhibited substantial clinical and methodological heterogeneity, with differences in study design, patient populations, healthcare settings, diagnostic methods, timing of microbiological investigations, study period, and local resource availability. This variability was reflected in the consistently high heterogeneity observed across analyses and limited the applicability of several meta-analytic approaches.

Publication bias was not formally assessed. Conventional methods, including funnel plots and Egger’s regression, are difficult to interpret in the presence of marked between-study heterogeneity and may yield misleading results. Consequently, the potential influence of publication bias cannot be excluded.

A further major source of heterogeneity was the absence of standardised reporting of microbiological outcomes. Even among studies with otherwise similar designs and patient populations, considerable variation was observed in definitions of bacterial co-infection, microbiological confirmation criteria, reporting practices, and outcome measures. These inconsistencies limit direct comparison of studies and reduced the precision of quantitative synthesis. Accordingly, the pooled estimates should be regarded as summary measures of a heterogeneous evidence base rather than precise estimates of the underlying prevalence of bacterial co-infection among patients with influenza.

Notwithstanding these limitations, this review provides a comprehensive and contemporary synthesis of the available evidence using rigorous systematic review and meta-analytic methods. In the absence of standardised reporting frameworks and sufficiently powered prospective studies using consistent microbiological definitions, such evidence syntheses remain important for identifying evidence gaps, guiding future research priorities, and informing clinical practice and antimicrobial stewardship strategies.

## Data Availability

All data produced in the present study are available upon reasonable request to the authors.

## Acknowledgements

The authors of this systematic review and meta-analyses would like to acknowledge the contributions of Dr. Paul Cannon in the development of the search strategy we used to undertake the review.

## Contribution

**Conceptualisation** - HB, AH. **Methodology** - HB, TS, AH. **Formal analysis** - HB. **Investigation** - HB, RP. **Data curation** – HB, RP. **Writing – Original Draft**. HB, AH. **Writing – Review & Editing** - All authors. **Visualisation**. HB. **Supervision.** TS, MEM, AH. **Funding acquisition** – TS, MEM, AH

