## Supplementary Material for "Bacterial co-infection and antimicrobial use in hospital-attended patients with laboratory-confirmed influenza infection: a systematic review and meta-analysis"

**SUPPLEMENTARY MATERIAL/APPENDIX**

**Table of Contents**

**Page 2 ~ Supplementary Table 1 –** Search strategy including search terms used in each corresponding database

**Page 6 ~** **Supplementary Table 2-5 –** Breakdown of microbiological testing

**Page 8 ~ Supplementary Table 6a & 6b –** GRADE assessments

**Page 10 ~ Supplementary Table 7** – Characteristics of included studies

**Page 17 ~** **Supplementary Table 8-12** – Joanna Briggs Institute critical appraisal results

**Page 27 ~** **Supplementary Table 13-19** – Bacterial species identification counts

**Page 31 ~ Supplementary Table 20 –** Excluded papers with reasons for exclusion

**Page 42 ~ Supplementary Table 21 –** Funding Information of included studies

**Page 52 ~ Supplementary Figure 1 -** Influence diagnostics using Cook’s distance

**Page 53 ~ Supplementary Figure 2** - Temporal distribution (Pandemic/Seasonal)

**Supplementary Table 1- Search strategy including search terms used in each corresponding database**

**MEDLINE**

1 exp Bacterial Infections/  
2 exp Pneumonia, Bacterial/  
3 bacteri*.ti,ab,kw.  
4 or/1-3  
5 exp Influenza, Human/ 
6 flu.ti,ab,kw.  
7 influenza*.ti,ab,kw.  
8 exp Influenza A virus/  
9 exp Influenza B virus/  
10 (h1n1 or h5n1 or h3n2).ti,ab,kw.  
11 or/5-10  
12 Epidemiologic studies/  
13 exp case control studies/  
14 exp cohort studies/ 
15 Case control.tw.  
16 (cohort adj (study or studies)).tw.  
17 Cohort analy$.tw.  
18 (Follow up adj (study or studies)).tw.  
19 (observational adj (study or studies)).tw.  
20 Longitudinal.tw.  
21 Retrospective.tw.  
22 Cross sectional.tw.  
23 Cross-sectional studies/  
24 or/12-23  
25 exp Coinfection/  
26 (coinfection* or co-infection* or mixed infection* or polymicrobial infection* or poly-microbial infection* or secondary infection*).ti,ab,kw.  
27 or/25-26  
28 Hospitals/  
29 hospital.tw.  
30 inpatient.tw.  
31 inpatient care.tw.  
32 secondary care.tw.  
33 acute care.tw.  
34 hospital setting.tw.  
35 hospital-based.tw.  
36 or/28-35  
37 4 and 11 and 24 and 27 and 36  
38 limit 37 to humans

**EMBASE** 

1 exp Bacterial Infection/  
2 exp bacterial pneumonia/ 
3 bacteri*.ti,ab,kw.  
4 or/1-3  
5 exp influenza/  
6 flu.ti,ab,kw.  
7 influenza.ti,ab,kw.  
8 exp influenza a/  
9 exp influenza b/  
10 (h1n1 or h5n1 or h3n2).ti,ab,kw.  
11 or/5-10  
12 exp epidemiology/  
13 exp case control studies/  
14 exp cohort studies/  
15 Case control.tw.  
16 (cohort adj (study or studies)).tw.  
17 Cohort analy$.tw.  
18 (observational adj (study or studies)).tw.  
19 Longitudinal.tw.  
20 Retrospective.tw.  
21 Cross sectional.tw.  
22 Cross-sectional studies/  
23 or/12-22  
24 exp coinfection/  
25 (coinfection* or co-infection* or mixed infection* or polymicrobial infection* or poly-microbial infection* or secondary infection*).ti,ab,kw.  
26 or/24-25  
27 exp hospitals/  
28 hospital.tw.  
29 inpatient.tw.  
30 inpatient care.tw.  
31 secondary care.tw.  
32 acute care.tw.  
33 hospital setting.tw.  
34 hospital-based.tw.  
35 or/27-34  
36 4 and 11 and 23 and 26 and 35  
37 limit 36 to humans

**Web of Science**

*Science Citation Index Expanded, Conference Proceedings Citation Index – Science, Emerging Sources Citation Index*

1 - TS=(bacteri* OR "bacterial infection*" OR "bacterial pneumonia") 
2 - TS=(influenza* OR flu OR "Influenza A virus" OR "Influenza B virus" OR h1n1 OR h5n1 OR h3n2) 
3 - TS=("epidemiologic studies" OR "case control" OR "case-control" OR "cohort study" OR "cohort studies" OR "cohort analy*" OR 
"follow up study" OR "follow-up study" OR "observational study" OR "longitudinal" OR "retrospective" OR 
"cross sectional" OR "cross-sectional study") 
4 - TS=("coinfection*" OR "co-infection*" OR "mixed infection*" OR "polymicrobial infection*" OR "poly-microbial infection*" OR "secondary infection*" OR "coinfection") 
5 - TS=("hospital" OR "inpatient" OR "inpatient care" OR "secondary care" OR "acute care" OR "hospital setting" OR "hospital-based") 
6 - #5 AND #4 AND #3 AND #2 AND #1

**SCOPUS**

( ( TITLE-ABS-KEY ( bacteri* ) OR INDEXTERMS ( "Bacterial Infections" ) OR INDEXTERMS ( "Pneumonia, Bacterial" ) ) ) AND ( TITLE-ABS-KEY ( flu OR influenza* OR h1n1 OR h5n1 OR h3n2 ) OR INDEXTERMS ( "Influenza, Human" ) OR INDEXTERMS ( "Influenza A virus" ) OR INDEXTERMS ( "Influenza B virus" ) ) AND ( TITLE-ABS-KEY ( "case control" OR ( cohort W/1 ( study OR studies ) ) OR "cohort analy*" OR ( "follow up" W/1 ( study OR studies ) ) OR ( "observational" W/1 ( study OR studies ) ) OR longitudinal OR retrospective OR "cross sectional" ) OR INDEXTERMS ( "Epidemiologic studies" ) OR INDEXTERMS ( "Case control studies" ) OR INDEXTERMS ( "Cohort studies" ) OR INDEXTERMS ( "Cross-sectional studies" ) ) AND ( TITLE-ABS-KEY ( coinfection* OR "co-infection*" OR "mixed infection*" OR "polymicrobial infection*" OR "poly-microbial infection*" OR "secondary infection*" ) OR INDEXTERMS ( "Coinfection" ) ) AND ( TITLE-ABS-KEY ( hospital OR inpatient OR "inpatient care" OR "secondary care" OR "acute care" OR "hospital setting" OR "hospital-based" ) OR INDEXTERMS ( "Hospitals" ) ) AND ( TITLE-ABS-KEY ( human* OR patient* OR adult* OR child* OR adolescent* ) )

PubMed

(

 "Bacterial Infections"[MeSH Terms] OR

 "Pneumonia, Bacterial"[MeSH Terms] OR

  bacteri*[tiab]

)

AND

(

  "Influenza, Human"[MeSH Terms] OR

  flu[tiab] OR

  influenza*[tiab] OR

  "Influenza A virus"[MeSH Terms] OR

  "Influenza B virus"[MeSH Terms] OR

  (h1n1[tiab] OR h5n1[tiab] OR h3n2[tiab])

)

AND

(

  "Epidemiologic Studies"[MeSH Terms] OR

  "Case-Control Studies"[MeSH Terms] OR

  "Cohort Studies"[MeSH Terms] OR

  "Cross-Sectional Studies"[MeSH Terms] OR

  "Observational Study"[Publication Type] OR

  "Retrospective Studies"[MeSH Terms] OR

  "Longitudinal Studies"[MeSH Terms] OR

  "Follow-Up Studies"[MeSH Terms] OR

  "case control"[tiab] OR

  (cohort[tiab] AND (study[tiab] OR studies[tiab])) OR

  "cohort analy*"[tiab] OR

  ("follow up"[tiab] AND (study[tiab] OR studies[tiab])) OR

  ("observational"[tiab] AND (study[tiab] OR studies[tiab])) OR

  "longitudinal"[tiab] OR

  "retrospective"[tiab] OR

  "cross sectional"[tiab]

)

AND

(

  "Coinfection"[MeSH Terms] OR

  coinfection*[tiab] OR

  co-infection*[tiab] OR

  mixed infection*[tiab] OR

  polymicrobial infection*[tiab] OR

  poly-microbial infection*[tiab] OR

  secondary infection*[tiab]

)

AND

(

  "Hospitals"[MeSH Terms] OR

  hospital[tiab] OR

  inpatient[tiab] OR

  "inpatient care"[tiab] OR

  "secondary care"[tiab] OR

  "acute care"[tiab] OR

  "hospital setting"[tiab] OR

  "hospital-based"[tiab]

)

AND

humans[MeSH Terms]

| **Supplementary Table 2 - Number of studies reporting each method of bacterial identification** | | | | |
| --- | --- | --- | --- | --- |
| **Method** | **Adults (n=56)** | **Paediatrics (n=29)** | **All Ages  (n=12)** | **Total (n=97)** |
| Bacterial Culture | 43 | 21 | 8 | 72 (74.2%) |
| Urinary Antigen Test | 19 | 4 | 3 | 26 (26.8%) |
| PCR | 8 | 4 | 3 | 15 (15.5%) |
| Unspecified | 12 | 6 | 1 | 19 (19.6%) |

| **Supplementary Table 3 - Number of studies reporting each bacterial sample method/sample site** | | | | |
| --- | --- | --- | --- | --- |
| **Method** | **Adults (n=56)** | **Paediatrics (n=29)** | **All Ages (n=12)** | **Total (n=97)** |
| Sputum | 32 | 8 | 7 | 47 (48.5%) |
| Bronchoalveolar lavage (BAL) | 23 | 8 | 5 | 36 (37.1%) |
| Tracheal | 14 | 3 | 4 | 21 (21.65%) |
| Pleural | 10 | 5 | 4 | 19 (19.6%) |
| Bronchial | 8 | 1 | 1 | 10 (10.3%) |
| Nasopharyngeal Swab/Aspirate | 6 | 9 | 1 | 16 (16.5%) |
| Serum/Blood | 34 | 18 | 8 | 60 (61.9%) |
| Urine | 21 | 3 | 4 | 28 (28.9%) |
| Unspecified | 12 | 5 | 1 | 18 (18.6%) |

| **Supplementary Table 4 - Number of studies reporting each method of Influenza testing** | | | | |
| --- | --- | --- | --- | --- |
| **Method** | **Adults (n=56)** | **Paediatrics (n=29)** | **All Ages (n=12)** | **Total (n=97)** |
| PCR | 48 | 24 | 11 | 83 (85.6%) |
| Antigen Testing | 6 | 13 | 3 | 22 (22.7%) |
| Viral Culture | 3 | 7 | 2 | 12 (12.4%) |
| Unspecified | 5 | 0 | 1 | 6 (6.2%) |

| **Supplementary Table 5 - Number of studies reporting each Influenza sample method/sample site** | | | | |
| --- | --- | --- | --- | --- |
| **Method** | **Adults (n=56)** | **Paediatrics (n=29)** | **Mixed (n=12)** | **Total (n=97)** |
| Nasopharyngeal Swab/Aspirate | 34 | 21 | 5 | 58 (59.8%) |
| Bronchoalveolar lavage (BAL) | 12 | 3 | 2 | 17 (17.5%) |
| Tracheal | 5 | 2 | 0 | 7 (7.2%) |
| Bronchial | 1 | 0 | 0 | 1 (1.0%) |
| Sputum | 4 | 3 | 1 | 8 (8.3%) |
| Serum/Blood | 3 | 4 | 0 | 7 (7.2%) |
| Unspecified | 19 | 5 | 7 | 31 (32.0%) |

| **Supplementary Table 6a - GRADE Assessment, Outcome 1.**  Pooled prevalence of bacterial co-infection | | |
| --- | --- | --- |
| **Outcome 1:** Pooled prevalence of bacterial co-infection | | |
| **Domain** | **Judgment** | **Reason for judgment** |
| **Starting certainty** | Low | Evidence is derived from Observational Studies |
| **Risk of bias** | Very serious | Predominantly (63.9%) retrospective study designs with non-systematic microbiological testing and potential for selection bias. |
| **Inconsistency** | Very serious | Substantial unexplained heterogeneity was observed (I² >96%), which was not resolved by subgroup analyses. |
| **Indirectness** | Very serious | Variability in diagnostic methods and timing of bacterial testing resulted in indirectness relative to the review question. |
| **Imprecision** | Moderate | While imprecision was not considered serious as the pooled estimate was based on a large total sample size (n = 116,273) high levels of heterogeneity and the variability of the 95% confidence intervals was noted. However, the findings were sufficiently consistent to support interpretation. |
| **Publication bias** | Not assessed | Publication bias was not formally assessed due to the nature of prevalence data. |
| **Other considerations (upgrade)** | None | No upgrading was applied as the outcome represents a prevalence estimate derived from observational data, for which GRADE upgrading criteria (large effect, dose–response, or residual confounding) are not applicable. |
| **Final certainty of evidence** | Very low | The overall certainty of evidence was judged to be very low due to serious risk of bias arising from non-systematic microbiological testing, serious inconsistency due to extreme unexplained heterogeneity, and serious indirectness related to variability in diagnostic methods and timing of bacterial testing. |

| **Supplementary Table 6b. GRADE Assessment - Outcome 2. Antimicrobial Therapy vs Microbiologically confirmed bacterial co-infection.** | | |
| --- | --- | --- |
| **Outcome 2:** Antimicrobial Therapy vs Microbiologically confirmed bacterial co-infection. | | |
| **Domain** | **Judgment** | **Reason for judgment** |
| **Starting certainty** | Low | Evidence is derived from Observational Studies |
| **Risk of bias** | Serious | The assessment concludes that there is a serious risk of bias due to predominantly observational study designs, clinician-driven and frequently empirical antimicrobial prescribing, broad and inconsistent definitions of antimicrobial exposure, and unclear temporal relationships between antibiotic initiation and microbiological testing. Most included studies were assessed as being at serious risk of bias. |
| **Inconsistency** | Serious | The assessment has found serious inconsistency due to extreme unexplained heterogeneity in antimicrobial prescribing practices across studies (I² = 99.86%), which was not reduced by subgroup analyses and likely reflects substantial variation in clinical practice, patient severity, and institutional prescribing policies. |
| **Indirectness** | Very serious | Variability in diagnostic methods and timing of bacterial testing resulted in indirectness relative to the review question. [Insert list of testing methods breakdown] |
| **Imprecision** | Moderate | Imprecision was not considered serious as the pooled estimate was based on a large total sample size (n = 58,910), with a high number of events, and despite variability, the 95% confidence intervals consistently indicated a high prevalence of antimicrobial use without spanning clinically divergent interpretations. |
| **Publication bias** | Not assessed | Publication bias was not formally assessed due to the descriptive nature of the outcome and substantial between-study heterogeneity; however, there was no clear indication of selective reporting. |
| **Other considerations (upgrade)** | None |  |
| **Final certainty of evidence** | Very low | The certainty of evidence was judged to be very low due to serious risk of bias related to clinician-driven empirical prescribing, serious inconsistency from extreme unexplained heterogeneity, and serious indirectness arising from incomplete microbiological testing and the inability to link antimicrobial use to confirmed bacterial co-infection at the individual patient level. |

| **Supplementary Table 7 – Characteristics of Included Studies (n=97)** | | | | | | | | | |
| --- | --- | --- | --- | --- | --- | --- | --- | --- | --- |
| **Study** | **Nation/Territory** | **Year** | **Age Group** | **Setting** | **Total Participants** | **Total Influenza Cases** | **Total Bacterial Co-infection Cases** | **Bacterial Co-infection Prevalence** | **% of Cohort Prescribed Antibiotics** |
| Abelenda-Alonso G, et al., 2020 | Spain | 2020 | Adults | Non-ICU (Hospital) | 1123 | 153 | 55 | 35.9% | - |
| Ahn S, et al., 2011 | South Korea | 2011 | Adults | Non-ICU (ED) | 96 | 60 | 16 | 26.7% | - |
| Al Ali A, et al. 2021 | UAE | 2021 | Paediatrics | Non-ICU (Hospital) | 1392 | 1392 | 165 | 11.9% | 37.0% |
| Al-Dorzi H.M, et al. 2024 | Saudi Arabia | 2024 | Adults | Non-ICU (ED) | 675 | 675 | 42 | 6.2% | 93.8% |
| Anania V.G, et al. 2020 | USA | 2020 | Paediatrics | ICU | 105 | 105 | 56 | 53.3% | - |
| Bartley P.S, et al. 2022 | USA | 2021 | Adults | Non-ICU (Hospital) | 4313 | 4313 | 445 | 10.3% | - |
| Bender J.M, et al. 2010 | USA | 2010 | Paediatrics | Non-ICU (Hospital) | 833 | 833 | 16 | 1.9% | - |
| Beumer M.C, et al. 2019 | Netherlands | 2019 | Mixed Cohort | Non-ICU (Hospital) | 199 | 199 | 26 | 13.1% | 44.7% |
| Bjarnason A, et al. 2012 | Iceland | 2012 | Adults | Non-ICU (Hospital) | 313 | 114 | 22 | 19.3% | 100.0% |
| Blyth C.C, et al. 2013 | Australia | 2012 | Adults | ICU | 198 | 198 | 36 | 18.2% | - |
| Carbonell R, et al. 2021 | Spain | 2021 | Adults | ICU | 1608 | 1608 | 422 | 26.2% | - |
| Chen J, et al. 2018 | China | 2018 | Paediatrics | Non-ICU (Hospital) | 1992 | 78 | 12 | 15.4% | - |
| Choi S.H, et al. 2015 | South Korea | 2015 | Adults | ICU | 78 | 51 | 19 | 37.3% | - |
| Christensen I, et al. 2023 | Norway | 2023 | Adults | Non-ICU (Hospital) | 116 | 116 | 17 | 14.7% | 76.0% |
| Cillóniz C, et al. 2012 | Spain | 2012 | Adults | Non-ICU (Hospital) | 667 | 128 | 42 | 32.8% | 100.0% |
| Cordero E, et al. 2012 | Spain | 2009 | Mixed Cohort | Non-ICU (Hospital) | 559 | 559 | 42 | 7.5% | 70.7% |
| Cuquemelle E, et al. 2011 | France | 2011 | Adults | ICU | 103 | 103 | 48 | 46.6% | - |
| Damasio G.A, et al. 2015 | Brazil | 2015 | Mixed Cohort | Non-ICU (Hospital) | 169 | 64 | 26 | 40.6% | 79.3% |
| Dawood F.S, et al. 2010 | USA | 2010 | Paediatrics | Non-ICU (Hospital) | 4015 | 4015 | 76 | 1.9% | - |
| Dhanoa A, et al. 2011 | Malaysia | 2011 | Mixed Cohort | Non-ICU (Hospital) | 50 | 50 | 14 | 28.0% | 82.0% |
| D'Onofrio V, et al. 2021 | Belgium | 2020 | Adults | Non-ICU (ED) | 213 | 103 | 11 | 10.7% | 76.7% |
| Dumas G, et al. 2023 | France | 2023 | Adults | ICU | 370 | 219 | 50 | 22.8% | 100.0% |
| Eşki A, et al. 2019 | Turkey | 2019 | Paediatrics | Non-ICU (Hospital) | 280 | 280 | 17 | 6.1% | 70.7% |
| Estenssoro E, et al. 2010 | Argentina | 2010 | Adults | ICU | 337 | 337 | 80 | 23.7% | 100.0% |
| Falsey A.R, et al. 2013 | USA | 2013 | Adults | Non-ICU (ED) | 842 | 90 | 10 | 11.1% | - |
| Feys S, et al. 2024 | Belgium | 2024 | Adults | ICU | 423 | 142 | 28 | 19.7% | 99.3% |
| Garg S, et al. 2015 | USA | 2015 | Adults | Non-ICU (Hospital) | 4765 | 4765 | 129 | 2.7% | - |
| Guo L, et al. 2019 | China | 2019 | Adults | Non-ICU (Hospital) | 528 | 242 | 59 | 24.4% | - |
| Gutiérrez-Pizarraya A, et al. 2012 | Spain | 2012 | Adults | Non-ICU (Hospital) | 130 | 130 | 9 | 6.9% | - |
| Haddara A, et al. 2024 | Lebanon | 2024 | Mixed Cohort | Non-ICU (Hospital) | 2049 | 2049 | 558 | 27.2% | 35.0% |
| Haeberer M, et al. 2024 | Spain | 2024 | Adults | Non-ICU (Hospital) | 1364 | 598 | 82 | 13.7% | - |
| Hagerman A, et al. 2015 | Switzerland | 2015 | Paediatrics | Non-ICU (Hospital) | 326 | 326 | 36 | 11.0% | 45.1% |
| Hall M.W, et al. 2013 | USA | 2013 | Paediatrics | ICU | 52 | 52 | 18 | 34.6% | - |
| Hayashi Y, et al. 2012 | Australia | 2011 | Adults | Non-ICU (Hospital) | 4491 | 4491 | 57 | 1.3% | - |
| Hedberg P, et al. 2022 | Sweden | 2022 | Adults | Non-ICU (ED) | 2238 | 775 | 209 | 27.0% | 83.9% |
| Hernandez-Bou S, et al. 2013 | Spain | 2013 | Paediatrics | Non-ICU (Hospital) | 308 | 308 | 29 | 9.4% | 64.6% |
| Hsing T, et al. 2022 | Taiwan | 2022 | Paediatrics | Non-ICU (Hospital) | 558 | 558 | 64 | 11.5% | - |
| Ishiguro T, et al. 2013 | Japan | 2013 | Adults | Non-ICU (Hospital) | 1032 | 97 | 36 | 37.1% | 100.0% |
| Jamoussi A, et al. 2022 | Tunisia | 2022 | Adults | ICU | 120 | 120 | 17 | 14.2% | 88.3% |
| Jorda A, et al. 2024 | Austria | 2023 | Adults | Non-ICU (Hospital) | 1337 | 180 | 20 | 11.1% | - |
| Khandaker G, et al. 2014 | Australia | 2014 | Paediatrics | Non-ICU (Hospital) | 506 | 506 | 29 | 5.7% | 54.0% |
| Kim H.S, et al. 2011 | South Korea | 2011 | Mixed Cohort | Non-ICU (Hospital) | 115 | 115 | 34 | 29.6% | - |
| Kim J.H, et al. 2018 | South Korea | 2018 | Mixed Cohort | Non-ICU (ED) | 244 | 244 | 102 | 41.8% | 97.1% |
| Kim S.H, et al. 2011 | South Korea | 2011 | Adults | ICU | 245 | 245 | 91 | 37.1% | - |
| Krammer M, et al. 2024 | Germany | 2024 | Mixed Cohort | Non-ICU (Hospital) | 16520 | 1346 | 52 | 5.4% | - |
| Kumar S, et al. 2010 | USA | 2010 | Paediatrics | Non-ICU (Hospital) | 75 | 75 | 1 | 1.3% | 80.0% |
| Kwon Y.S, et al. 2017 | South Korea | 2017 | Adults | Non-ICU (Hospital) | 399 | 312 | 32 | 10.3% | - |
| Le Glass E, et al. 2022 | France | 2022 | Adults | Non-ICU (Hospital) | 4059 | 4059 | 166 | 4.1% | - |
| Lee W.C, et al. 2022 | Taiwan | 2022 | Adults | ICU | 117 | 117 | 32 | 27.4% | - |
| Li Y.N, et al. 2025 | China | 2025 | Paediatrics | Non-ICU (ED) | 39110 | 39110 | 910 | 2.3% | 13.0% |
| Li Z, et al. 2019 | China | 2019 | Paediatrics | Non-ICU (Hospital) | 3180 | 3180 | 226 | 7.1% | - |
| Liderot K, et al. 2013 | Sweden | 2013 | Adults | Non-ICU (Hospital) | 1094 | 1094 | 71 | 6.5% | 4.7% |
| Liu J, et al. 2020 | China | 2020 | Paediatrics | Non-ICU (Hospital) | 10429 | 1720 | 1140 | 66.3% | - |
| Liu Y, et al. 2021 | Hong Kong | 2021 | Adults | Non-ICU (Hospital) | 12160 | 12160 | 538 | 4.4% | - |
| Lopez-Delgado J.C, et al. 2013 | Spain | 2013 | Adults | ICU | 60 | 60 | 10 | 16.7% | 100.0% |
| Lu Y, et al. 2013 | China | 2013 | Paediatrics | Non-ICU (Hospital) | 720 | 95 | 14 | 14.7% | - |
| Mangas-Moro A, et al. 2023 | Spain | 2023 | Adults | Non-ICU (Hospital) | 1260 | 1260 | 436 | 34.6% | - |
| Marin-Corral J, et al. 2018 | Spain | 2018 | Adults | ICU | 2205 | 2205 | 383 | 17.37% | - |
| Mercat A, et al. 2011 | France | 2011 | Adults | ICU | 562 | 562 | 127 | 22.60% | - |
| Marcoux D, et al. 2022 | Belgium | 2022 | Adults | ICU | 112 | 55 | 40 | 72.7% | 39.3% |
| Martin-Loeches I, et al 2017 | Spain | 2016 | Adults | ICU | 2901 | 2901 | 482 | 16.6% | - |
| Melamed K.H, et al. 2020 | USA | 2020 | Adults | Non-ICU (Hospital) | 134 | 57 | 11 | 19.0% | - |
| Mendoza M.A, et al. 2023 | USA | 2022 | Adults | Non-ICU (Hospital) | 377 | 100 | 19 | 19.0% | - |
| Merișescu M.M, et al. 2023 | Romania | 2023 | Paediatrics | Non-ICU (Hospital) | 301 | 301 | 26 | 8.6% | - |
| Muscedere J, et al. 2013 | Canada | 2013 | Adults | ICU | 681 | 583 | 154 | 26.4% | 97.1% |
| Nguyen T, et al. 2012 | USA | 2012 | Paediatrics | ICU | 66 | 66 | 34 | 51.5% | - |
| Nolan V.G, et al. 2018 | USA | 2018 | Paediatrics | Non-ICU (Hospital) | 2219 | 149 | 13 | 8.7% | 88.0% |
| Owayed A.F, et al. 2012 | Kuwait | 2011 | Paediatrics | Non-ICU (Hospital) | 197 | 197 | 3 | 1.5% | 74.6% |
| Poulakou G, et al. 2011 | Spain | 2012 | Mixed Cohort | ICU | 53 | 53 | 8 | 15.1% | 58.9% |
| Qin T, et al. 2020 | China | 2020 | Mixed Cohort | Non-ICU (Hospital) | 52 | 52 | 24 | 46.2% | - |
| Randolph A.G, et al. 2011 | USA | 2011 | Paediatrics | ICU | 838 | 838 | 274 | 32.7% | - |
| Rice T.W, et al. 2012 | USA | 2012 | Mixed Cohort | ICU | 683 | 683 | 255 | 37.3% | - |
| Rouzé A, et al. 2021 | Pan-Europe | 2021 | Adults | ICU | 1050 | 482 | 162 | 33.6% | 88.5% |
| Rozencwajg S, et al. 2018 | France | 2018 | Adults | ICU | 77 | 77 | 39 | 50.6% | - |
| Schoettler J.J, et al. 2023 | Germany | 2023 | Adults | ICU | 190 | 76 | 5 | 6.6% | - |
| Shafran N, et al. 2021 | Israel | 2021 | Adults | Non-ICU (Hospital) | 1366 | 724 | 63 | 8.7% | - |
| Shah NS, et al. 2016 | USA | 2016 | Mixed Cohort | ICU | 507 | 507 | 114 | 22.5% | 92.1% |
| Shi T, et al. 2019 | China | 2019 | Paediatrics | ICU | 77 | 77 | 28 | 36.4% | 84.4% |
| Sohn C.H, et al. 2013 | South Korea | 2013 | Adults | Non-ICU (Hospital) | 135 | 59 | 15 | 25.4% | - |
| Soler-Font M, et al. 2022 | Spain | 2022 | Adults | Non-ICU (Hospital) | 2742 | 2742 | 603 | 22.0% | - |
| Tao RJ, et al. 2018 | China | 2018 | Adults | Non-ICU (Hospital) | 320 | 57 | 18 | 31.6% | - |
| Tasar S, et al. 2022 | Turkey | 2022 | Paediatrics | ICU | 145 | 74 | 15 | 20.3% | - |
| Teng F, et al. 2019 | China | 2019 | Adults | Non-ICU (Hospital) | 209 | 168 | 41 | 24.4% | - |
| Thelen J.M, et al. 2021 | Netherlands | 2021 | Adults | Non-ICU (ED) | 1635 | 653 | 23 | 3.5% | - |
| Tian J, et al. 2023 | China | 2023 | Adults | Non-ICU (Hospital) | 203 | 129 | 39 | 30.2% | - |
| Tokuhira N, et al. 2012 | Japan | 2012 | Paediatrics | ICU | 81 | 81 | 8 | 9.9% | 87.7% |
| Tsai C.F, et al. 2023 | Taiwan | 2023 | Paediatrics | Non-ICU (Hospital) | 1030 | 1030 | 161 | 15.6% | - |
| Üzüm Ö, et al. 2022 | Turkey | 2022 | Paediatrics | Non-ICU (Hospital) | 93 | 93 | 53 | 57.0% | 56.9% |
| Verdier V, et al. 2023 | Réunion | 2023 | Adults | ICU | 379 | 350 | 116 | 43.7% | - |
| Viasus D, et al. 2011 | Spain | 2011 | Adults | Non-ICU (Hospital) | 585 | 585 | 45 | 7.7% | 71.7% |
| Viasus D, et al. 2013 | Spain | 2013 | Adults | Non-ICU (Hospital) | 747 | 101 | 14 | 13.9% | - |
| von Baum H, et al. 2011 | Germany | 2011 | Adults | Non-ICU (Hospital) | 5032 | 160 | 34 | 21.3% | - |
| Wallemacq S, et al. 2022 | Belgium | 2022 | Adults | Non-ICU (Hospital) | 1610 | 1610 | 104 | 6.5% | - |
| Wei L, et al. 2015 | China | 2015 | Paediatrics | Non-ICU (Hospital) | 3181 | 551 | 121 | 22.0% | - |
| Zhang Q, et al. 2011 | China | 2011 | Paediatrics | Non-ICU (Hospital) | 821 | 75 | 15 | 20.0% | - |
| Zhang Y, et al. 2020 | China | 2020 | Adults | Non-ICU (Hospital) | 330 | 279 | 112 | 40.1% | 84.2% |
| Zhong PP, et al. 2016 | China | 2016 | Paediatrics | Non-ICU (Hospital) | 357 | 357 | 79 | 22.13% | - |

| **Supplementary Table 8 – Joanna Briggs Institute Appraisal, Cohort studies.** | | | | | | | |
| --- | --- | --- | --- | --- | --- | --- | --- |
| **Study** | **1 - Selection and Allocation** | **2 - Classification of Exposure** | **3 - Confounding Factors** | **4 - Temporal Precedence** | **5 - Assessment, Detection & Measurement** | **6 - Retention & Follow up** | **7 - Statistical Analyses** |
| Abelenda-Alonso G, et al., 2020 | N/A | YES | UNCLEAR | NO | YES | N/A | YES |
| Al Ali A, et al. 2021 | N/A | YES | YES | NO | YES | N/A | YES |
| Al-Dorzi H.M, et al. 2024 | YES | YES | UNCLEAR | NO | YES | N/A | YES |
| Anania V.G, et al. 2020 | N/A | YES | NO | NO | YES | N/A | YES |
| Bartley P.S, et al. 2022 | YES | YES | YES | UNCLEAR | YES | N/A |  |
| Bender J.M, et al. 2010 | N/A | YES | UNCLEAR | UNCLEAR | YES | N/A | YES |
| Beumer M.C, et al. 2019 | N/A | YES | UNCLEAR | NO | YES | N/A | YES |
| Bjarnason A, et al. 2012 | N/A | YES | NO | NO | YES | N/A |  |
| Carbonell R, et al. 2021 | N/A | UNCLEAR | UNCLEAR | NO | YES | N/A | YES |
| Choi S.H, et al. 2015 | YES | YES | NO | UNCLEAR | YES | N/A | YES |
| Christensen I, et al. 2023 | N/A | YES | YES | UNCLEAR | YES | N/A |  |
| Cillóniz C, et al. 2012 | N/A | YES | YES | NO | YES | N/A | YES |
| Cordero E, et al. 2012 | YES | YES | NO | UNCLEAR | YES | YES | YES |
| D'Onofrio V, et al. 2021 | YES | YES | NO | NO | YES | UNCLEAR | YES |
| Damasio G.A, et al. 2015 | N/A | YES | YES | NO | YES | N/A | YES |
| Dhanoa A, et al. 2011 | N/A | YES | NO | NO | YES | N/A | YES |
| Dumas G, et al. 2023 | YES | YES | UNCLEAR | NO | YES | N/A | YES |
| Eşki A, et al. 2019 | N/A | YES | NO | NO | YES | NO | YES |
| Estenssoro E, et al. 2010 | N/A | YES | NO | NO | YES | N/A | YES |
| Falsey A.R, et al. 2013 | N/A | YES | YES | NO | YES | N/A | YES |
| Feys S, et al. 2024 | YES | YES | YES | NO | YES | N/A | YES |
| Garg S, et al. 2015 | N/A | YES | UNCLEAR | NO | YES | N/A | YES |
| Guo L, et al. 2019 | YES | YES | NO | NO | YES | N/A | YES |
| Gutiérrez-Pizarraya A, et al. 2012 | N/A | UNCLEAR | UNCLEAR | UNCLEAR | UNCLEAR | N/A | YES |
| Haddara A, et al. 2024 | N/A | YES | UNCLEAR | NO | YES | N/A | YES |
| Haeberer M, et al. 2024 | YES | YES | YES | NO | YES | N/A |  |
| Hagerman A, et al. 2015 | N/A | YES | NO | NO | YES | N/A | YES |
| Hall M.W, et al. 2013 | YES | YES | UNCLEAR | UNCLEAR | YES | N/A | YES |
| Hedberg P, et al. 2022 | YES | YES | UNCLEAR | NO | YES | N/A | YES |
| Ishiguro T, et al. 2013 | N/A | YES | NO | UNCLEAR | YES | N/A | YES |
| Jamoussi A, et al. 2022 | N/A | YES | NO | NO | YES | N/A | YES |
| Jorda A, et al. 2024 | YES | YES | NO | NO | YES | N/A | YES |
| Khandaker G, et al. 2014 | N/A | YES | YES | NO | YES | N/A |  |
| Kim J.H, et al. 2018 | N/A | YES | YES | UNCLEAR | YES | N/A | YES |
| Kim S.H, et al. 2011 | YES | YES | YES | NO | YES | N/A | YES |
| Kumar S, et al. 2010 | N/A | YES | NO | NO | YES | N/A | YES |
| Kwon Y.S, et al. 2017 | YES | YES | YES | NO | YES | N/A | YES |
| Le Glass E, et al. 2022 | YES | YES | NO | UNCLEAR | YES | N/A |  |
| Li Z, et al. 2019 | YES | YES | NO | NO | YES | N/A | YES |
| Liderot K, et al. 2013 | YES | YES | NO | NO | YES | N/A | YES |
| Liu Y, et al. 2021 | YES | YES | YES | NO | YES | N/A |  |
| Lopez-Delgado J.C, et al. 2013 | NO | YES | YES | NO | YES | N/A | YES |
| Marcoux D, et al. 2022 | YES | YES | NO | NO | YES | N/A |  |
| Marin-Corral J, et al. 2018 | UNCLEAR | YES | UNCLEAR | UNCLEAR | YES | N/A | YES |
| Martin-Loeches I, et al 2017 | NO | YES | NO | UNCLEAR | YES | N/A | YES |
| Melamed K.H, et al. 2020 | YES | YES | YES | NO | YES | YES | YES |
| Mendoza M.A, et al. 2023 | YES | YES | NO | NO | YES | N/A | YES |
| Mercat A, et al. 2011 | UNCLEAR | YES | UNCLEAR | UNCLEAR | YES | N/A | N/A |
| Muscedere J, et al. 2013 | N/A | YES | UNCLEAR | NO | YES | N/A | YES |
| Nguyen T, et al. 2012 | NO | YES | UNCLEAR | NO | YES | YES | YES |
| Nolan V.G, et al. 2018 | NO | YES | UNCLEAR | NO | YES | YES | YES |
| Poulakou G, et al. 2011 | N/A | YES | NO | NO | YES | N/A | YES |
| Randolph A.G, et al. 2011 | N/A | UNCLEAR | NO | UNCLEAR | YES | N/A | YES |
| Rice T.W, et al. 2012 | YES | YES | UNCLEAR | NO | YES | N/A | YES |
| Rouzé A, et al. 2021 | YES | YES | YES | NO | YES | N/A | YES |
| Rozencwajg S, et al. 2018 | N/A | YES | YES | NO | YES | N/A | YES |
| Shafran N, et al. 2021 | YES | YES | UNCLEAR | NO | YES | N/A | YES |
| Shah NS, et al. 2016 | YES | YES | UNCLEAR | NO | YES | N/A | YES |
| Shi T, et al. 2019 | N/A | YES | NO | NO | YES | N/A | YES |
| Sohn C.H, et al. 2013 | YES | YES | UNCLEAR | NO | YES | N/A | YES |
| Tao RJ, et al. 2018 | YES | YES | NO | UNCLEAR | YES | N/A | YES |
| Tasar S, et al. 2022 | YES | YES | NO | NO | YES | N/A | YES |
| Teng F, et al. 2019 | N/A | YES | YES | NO | YES | N/A | YES |
| Thelen J.M, et al. 2021 | YES | YES | NO | NO | YES | N/A | YES |
| Tian J, et al. 2023 | YES | YES | NO | NO | YES | N/A |  |
| Tsai C.F, et al. 2023 | YES | YES | NO | NO | YES | N/A | YES |
| Verdier V, et al. 2023 | N/A | YES | NO | NO | YES | N/A | YES |
| Viasus D, et al. 2011 | N/A | YES | NO | NO | YES | YES | YES |
| Viasus D, et al. 2013 | YES | YES | NO | UNCLEAR | YES | N/A | YES |
| von Baum H, et al. 2011 | NO | YES | NO | NO | YES | NO | YES |
| Wallemacq S, et al. 2022 | YES | YES | NO | NO | YES | N/A |  |
| Wei L, et al. 2015 | YES | YES | NO | NO | YES | YES | YES |
| Zhang Q, et al. 2011 | UNCLEAR | YES | NO | NO | YES | NO | YES |
| Zhang Y, et al. 2020 | YES | YES | NO | NO | YES | N/A | YES |

| **Supplementary Table 9 – Joanna Briggs Institute Appraisal, Cross-sectional studies.**  1 - Were the criteria for inclusion in the sample clearly defined?  2 -Were objective, standard criteria used for measurement of the condition?  3 - Was the exposure measured in a valid and reliable way?  4 - Were the outcomes measured in a valid and reliable way?  5 - Were confounding factors identified?  6 - Were strategies to deal with confounding factors stated?  7 - Was appropriate statistical analysis used? | | | | | | | | |
| --- | --- | --- | --- | --- | --- | --- | --- | --- |
| **Study_ID** | **1** | **2** | **3** | **4** | **5** | **6** | **7** | **8** |
| Li Y.N, et al. 2025 | YES | UNCLEAR | YES | YES | YES | YES | YES | YES |
| Soler-Font M, et al. 2022 | YES | YES | UNCLEAR | UNCLEAR | NO | NO | UNCLEAR | YES |
| Üzüm Ö, et al. 2022 | YES | YES | YES | YES | NO | NO | UNCLEAR | YES |
| Chen J, et al. 2018 | YES | YES | YES | YES | NO | NO | UNCLEAR | YES |
| Mangas-Moro A, et al. 2023 | YES | YES | YES | YES | NO | NO | YES | YES |

| **Supplementary Table 10 – Joanna Briggs Institute Appraisal, Prevalence studies.**  1 - Was the sample frame appropriate to address the target population?  2 - Were study participants sampled in an appropriate way?  3 - Was the sample size adequate?  4 - Were the study subjects and the setting described in detail?  6 - Were valid methods used for the identification of the condition?  7 - Was the condition measured in a standard, reliable way for all participants?  8 - Was there appropriate statistical analysis?  9 - Was the response rate adequate, and if not, was the low response rate managed appropriately? | | | | | | | | | |
| --- | --- | --- | --- | --- | --- | --- | --- | --- | --- |
| **Study** | **1** | **2** | **3** | **4** | **5** | **6** | **7** | **8** | **9** |
| Krammer M, et al. 2024 | YES | YES | YES | UNCLEAR | YES | YES | NO | YES | YES |
| Hsing T, et al. 2022 | YES | YES | YES | YES | YES | YES | NO | YES | YES |
| Hayashi Y, et al. 2012 | YES | YES | YES | UNCLEAR | UNCLEAR | YES | NO | YES | YES |
| Blyth C.C, et al. 2013 | YES | YES | UNCLEAR | YES | YES | YES | NO | YES | YES |

| **Supplementary Table 11 – Joanna Briggs Institute Appraisal, Case Series studies.**  1 - Were there clear criteria for inclusion in the case series?   2 - Was the condition measured in a standard, reliable way for all participants included in the case series?  3 - Were valid methods used for identification of the condition for all participants included in the case series?   4 - Did the case series have consecutive inclusion of participants?   5 - Did the case series have complete inclusion of participants?   6 - Was there clear reporting of the demographics of the participants in the study?   7 - Was there clear reporting of clinical information of the participants?   8 - Were the outcomes or follow up results of cases clearly reported?  9 - Was there clear reporting of the presenting site(s)/clinic(s) demographic information?  10 - Was statistical analysis appropriate? | | | | | | | | | | |
| --- | --- | --- | --- | --- | --- | --- | --- | --- | --- | --- |
| Study | 1 | 2 | 3 | 4 | 5 | 6 | 7 | 8 | 9 | 10 |
| Merișescu M.M, et al. 2023 | YES | YES | YES | YES | UNCLEAR | YES | YES | YES | UNCLEAR | YES |
| Lee W.C, et al. 2022 | YES | YES | YES | UNCLEAR | UNCLEAR | YES | YES | YES | YES | YES |
| Owayed A.F, et al. 2012 | YES | UNCLEAR | UNCLEAR | UNCLEAR | NO | YES | YES | NO | YES | UNCLEAR |
| Dawood F.S, et al. 2010 | YES | YES | YES | YES | UNCLEAR | YES | YES | YES | YES | UNCLEAR |
| Hernandez-Bou S, et al. 2013 | YES | YES | UNCLEAR | UNCLEAR | UNCLEAR | NO | NO | NO | NO | UNCLEAR |
| Tokuhira N, et al. 2012 | YES | YES | UNCLEAR | YES | UNCLEAR | UNCLEAR | UNCLEAR | UNCLEAR | UNCLEAR | UNCLEAR |
| Cuquemelle E, et al. 2011 | YES | YES | YES | UNCLEAR | NO | YES | YES | UNCLEAR | UNCLEAR | YES |
| Kim H.S, et al. 2011 | YES | YES | YES | UNCLEAR | UNCLEAR | YES | YES | YES | UNCLEAR | YES |
| Ahn S, et al., 2011 | YES | UNCLEAR | YES | UNCLEAR | UNCLEAR | YES | YES | UNCLEAR | UNCLEAR | YES |
| Lu Y, et al. 2013 | YES | YES | YES | UNCLEAR | UNCLEAR | UNCLEAR | YES | YES | YES | YES |
| Zhong PP, et al. 2016 | YES | YES | YES | UNCLEAR | UNCLEAR | UNCLEAR | YES | YES | NO | UNCLEAR |

| **Supplementary Table 12 – Joanna Briggs Institute Appraisal, Case Series studies.**  1 - Were the groups comparable other than the presence of disease in cases or the absence of disease in controls?   2 - Were cases and controls matched appropriately?  3 - Were the same criteria used for identification of cases and controls?  4 - Was exposure measured in a standard, valid and reliable way?  5 - Was exposure measured in the same way for cases and controls?  6 - Were confounding factors  7 - Were strategies to deal with confounding factors stated? identified?  8 - Were outcomes assessed in a standard, valid and reliable way for cases and controls?  9 - Was the exposure period of interest long enough to be meaningful?  10 - Was appropriate statistical analysis used? | | | | | | | | | | |
| --- | --- | --- | --- | --- | --- | --- | --- | --- | --- | --- |
| **Study** | **1** | **2** | **3** | **4** | **5** | **6** | **7** | **8** | **9** | **10** |
| Schoettler J.J, et al. 2023 | NO | NO | UNCLEAR | UNCLEAR | UNCLEAR | YES | NO | YES | UNCLEAR | YES |
| Liu J, et al. 2020 | YES | UNCLEAR | YES | UNCLEAR | YES | NO | NO | YES | UNCLEAR | YES |
| Qin T, et al. 2020 | NO | NO | UNCLEAR | UNCLEAR | YES | NO | NO | YES | UNCLEAR | YES |

| **Supplementary Table 13 – Bacterial Species Identification counts, Adults subgroup (n=35)** | | |
| --- | --- | --- |
| **Species** | **n** | **%** |
| *Acinetobacter baumannii* | 80 | 3.41% |
| *Aspergillus fumigatus* | 39 | 1.66% |
| *Escherichia coli* | 65 | 2.77% |
| *Haemophilus influenzae* | 191 | 8.15% |
| *Klebsiella pneumoniae* | 89 | 3.80% |
| *Moraxella catarrhalis* | 28 | 1.19% |
| *Mycoplasma pneumoniae* | 13 | 0.55% |
| *Pseudomonas aeruginosa* | 190 | 8.11% |
| *Staphylococcus aureus* | 648 | 27.65% |
| *Streptococcus pneumoniae* | 927 | 39.55% |
| Other | 74 | 3.16% |
| **Total** | 2344 |  |

| **Supplementary Table 14 – Bacterial Species Identification counts, Paediatrics subgroup (n=15)** | | |
| --- | --- | --- |
| **Species** | **n** | **%** |
| *Acinetobacter baumannii* | 3 | 0.49% |
| *Aspergillus fumigatus* | 0 | 0.00% |
| *Escherichia coli* | 24 | 3.90% |
| *Haemophilus influenzae* | 64 | 10.41% |
| *Klebsiella pneumoniae* | 17 | 2.76% |
| *Moraxella catarrhalis* | 28 | 4.55% |
| *Mycoplasma pneumoniae* | 10 | 1.63% |
| *Pseudomonas aeruginosa* | 16 | 2.60% |
| *Staphylococcus aureus* | 238 | 38.70% |
| *Streptococcus pneumoniae* | 185 | 30.08% |
| *Other* | 30 | 4.88% |
| **Total** | 615 |  |

| **Supplementary Table 15 – Bacterial Species Identification counts, All Ages (n=59)** | | |
| --- | --- | --- |
| **Species** | **n** | **%** |
| *Acinetobacter baumannii* | 88 | 2.60% |
| *Aspergillus fumigatus* | 39 | 1.15% |
| *Escherichia coli* | 89 | 2.63% |
| *Haemophilus influenzae* | 275 | 8.14% |
| *Klebsiella pneumoniae* | 116 | 3.43% |
| *Moraxella catarrhalis* | 63 | 1.86% |
| *Mycoplasma pneumoniae* | 69 | 2.04% |
| *Pseudomonas aeruginosa* | 227 | 6.72% |
| *Staphylococcus aureus* | 1012 | 29.95% |
| *Streptococcus pneumoniae* | 1191 | 35.25% |
| *Other* | 122 | 3.61% |
| **Total** | 3379 |  |

| **Supplementary Table 16 – Bacterial Species Identification counts, ICU subgroup (n=23)** | | |
| --- | --- | --- |
| **Species** | **n** | **%** |
| *Acinetobacter baumannii* | 57 | 3.72% |
| *Aspergillus fumigatus* | 39 | 2.54% |
| *Escherichia coli* | 23 | 1.50% |
| *Haemophilus influenzae* | 94 | 6.13% |
| *Klebsiella pneumoniae* | 53 | 3.46% |
| *Moraxella catarrhalis* | 16 | 1.04% |
| *Mycoplasma pneumoniae* | 1 | 0.07% |
| *Pseudomonas aeruginosa* | 121 | 7.89% |
| *Staphylococcus aureus* | 529 | 34.51% |
| *Streptococcus pneumoniae* | 506 | 33.01% |
| *Other* | 94 | 6.13% |
| **Total** | 1533 |  |

| **Supplementary Table 17 – Bacterial Species Identification counts, Non-ICU subgroup (n=36)** | | |
| --- | --- | --- |
| **Species** | **n** | **%** |
| *Acinetobacter baumannii* | 31 | 1.72% |
| *Aspergillus fumigatus* | 0 | 0.00% |
| *Escherichia coli* | 66 | 3.66% |
| *Haemophilus influenzae* | 181 | 10.04% |
| *Klebsiella pneumoniae* | 63 | 3.50% |
| *Moraxella catarrhalis* | 47 | 2.61% |
| *Mycoplasma pneumoniae* | 68 | 3.77% |
| *Pseudomonas aeruginosa* | 106 | 5.88% |
| *Staphylococcus aureus* | 483 | 26.80% |
| *Streptococcus pneumoniae* | 685 | 38.01% |
| *Other* | 72 | 4.00% |
| **Total** | 1802 |  |

| **Supplementary Table 18 – Bacterial Species Identification counts, Pandemic subgroup (n=44)** | | |
| --- | --- | --- |
| **Species** | **n** | **%** |
| *Acinetobacter baumannii* | 62 | 2.26% |
| *Aspergillus fumigatus* | 65 | 2.37% |
| *Escherichia coli* | 65 | 2.37% |
| *Haemophilus influenzae* | 214 | 7.81% |
| *Klebsiella pneumoniae* | 115 | 4.20% |
| *Moraxella catarrhalis* | 48 | 1.75% |
| *Mycoplasma pneumoniae* | 61 | 2.23% |
| *Pseudomonas aeruginosa* | 197 | 7.19% |
| *Staphylococcus aureus* | 803 | 29.32% |
| *Streptococcus pneumoniae* | 983 | 35.89% |
| *Other* | 126 | 4.60% |
| **Total** | 2739 |  |

| **Supplementary Table 19 – Bacterial Species Identification counts, Seasonal subgroup (n=15)** | | |
| --- | --- | --- |
| **Species** | **n** | **%** |
| *Acinetobacter baumannii* | 0 | 0.00% |
| *Aspergillus fumigatus* | 0 | 0.00% |
| *Escherichia coli* | 24 | 4.03% |
| *Haemophilus influenzae* | 61 | 10.23% |
| *Klebsiella pneumoniae* | 1 | 0.17% |
| *Moraxella catarrhalis* | 15 | 2.52% |
| *Mycoplasma pneumoniae* | 8 | 1.34% |
| *Pseudomonas aeruginosa* | 30 | 5.03% |
| *Staphylococcus aureus* | 209 | 35.07% |
| *Streptococcus pneumoniae* | 208 | 34.90% |
| *Other* | 40 | 6.71% |
| **Total** | 596 |  |

| **Supplementary Table 20 – Excluded Papers with reasons for exclusion.** | | | |
| --- | --- | --- | --- |
| Paper | Title | Reason for exclusion | Notes |
| [Unknown Authors] | The Effectiveness of Adding Multiple Intermittent High-dose Inhalations of Nitric Oxide to Standard Antibacterial Therapy in the Treatment of PneumoniaA1 - Anonymous. | Ineligible Study Design | Clinical trial |
| [Unknown Authors] | Cohort Study Evaluating the Clinical Effectiveness, Safety and Immunogenicity to the Pandemic Influenza Vaccination in Patients With Cystic Fibrosis and, Where Applicable, the Clinical Expression of Influenza A (H1N1)A1 - Anonymous. | Ineligible Study Design | Clinical trial |
| [Unknown Authors] | Procalcitonin Role in Influenza Patients With Regard to Morbidity, Mortality and Antibiotic UseA1 - Anonymous. | Ineligible Study Design | Clinical trial |
| [Unknown Authors] | Pneumonia in Children: Aetiology, Ideal Antibiotic Duration, Quality of LifeA1 - Anonymous. | Ineligible Study Design | Clinical trial |
| Ahn, S. et al.  2010 | Procalcitonin in 2009 H1N1 influenza pneumonia: Role in differentiating from bacterial pneumonia | Non-English Full Text | Korean Full Text |
| Antunes, A.P. et al. 2019 | INFLUENZA A - A CALM AFTER THE STORM? | Abstract Only |  |
| Arjarquah, AK. et al 2022 | Occurrence of influenza and bacterial infections in cancer patients receiving radiotherapy in Ghana | <50 Influenza Cases |  |
| Aston, SJ. et al. 2019 | Etiology and Risk Factors for Mortality in an Adult Community-acquired Pneumonia Cohort in Malawi | <50 Influenza Cases |  |
| Aquino-Esperanza, J. et al. 2010 | Severe respiratory disease in an intensive care unit during influenza A(H1N1)2009 pandemia | No Access | No DOI |
| Baird, J.S. et al 2012 | Comparing the clinical severity of the first versus second wave of 2009 Influenza A (H1N1) in a New York City pediatric healthcare facility | Insufficient Data on Bacterial Coinfection |  |
| Bal, A. et al 2020 | Influenza -induced acute respiratory distress syndrome during the 2010-2016 seasons: bacterial co -infections and outcomes by virus type and subtype | <50 Influenza Cases |  |
| Barbadillo S, et al. 2016 | Clinical differences between influenza A (H1N1)PDM09 and influenza A(H3N2) infection in critically ill patients | Abstract Only |  |
| Bartley P, et al. 2019 | Influenza and bacterial pneumonia coinfection: Rates and outcomes | Abstract Only |  |
| Bello S, et al. 2014 | Inflammatory response in mixed viral-bacterial community-acquired pneumonia | <50 Influenza Cases |  |
| Beumer, CM. et al. 2017 | Influenza-associated risk factors for ICU admission and mortality | Abstract Only |  |
| Bhat, N. et al. 2005 | Influenza-associated deaths among children in the United States, 2003-2004 | Ineligible Study Design | Not all Influenza cases confirmed |
| Bouziri, A. et al 2016 | Severe acute respiratory infections caused by influenza a (H1N1)PDM09 in a pediatric intensive care unit during the 2015-16 season | <50 Influenza Cases |  |
| Brebner, J. et al 2011 | Pneumonia in H1N1 influenza infection | Abstract Only |  |
| Brown, M. et al 2024 | Purpura fulminans in young women with influenza and co-infections | Ineligible Patient Population | Purpura Fulminans |
| Cacacho, ALF. et al 2016 | H1N1pdm09 is an independent risk factor for severe influenza | Abstract Only |  |
| Cantais, A. et al 2014 | Epidemiology and microbiological investigations of community-acquired pneumonia in children admitted at the emergency department of a university hospital | <50 Influenza Cases |  |
| Chavan, R.D. et al 2015 | Surveillance of acute respiratory infections in Mumbai during 2011-12 | <50 Influenza Cases |  |
| Chen WC. et al 2017 | Bacterial coinfection in pandemic 2016 severe complicated influenza in Taiwan | Abstract Only |  |
| Chen YF. et al 2017 | Risk factors and outcomes of severe influenza pneumonia with bacterial co-infection in critically ill patients | Abstract Only |  |
| Chertow, DS. et al 2012 | Contribution of bacterial coinfection to severe influenza infection | Abstract Only | Editorial |
| Chien, YS. et al 2018 | Early corticosteroid in influenza pneumonia related acute respiratory distress syndrome treated with extracorporeal membrane oxygenation | Abstract Only |  |
| Choi SH. et al. 2012 | Viral infection in patients with severe pneumonia requiring intensive care unit admission | <50 Influenza Cases |  |
| Chong, LL. et al 2017 | Influenza and bacterial co-infections: The gold coast hospital and health service (GCHHS) experience | Abstract Only |  |
| Chuaychoo, B. et al 2021 | Characteristics, complications, and mortality of respiratory syncytial virus compared with influenza infections in hospitalized adult patients in Thailand | Insufficient Data on Bacterial Coinfection |  |
| Chu, RBH. et al 2023 | Comparison of COVID-19 with influenza A in the ICU: a territory-wide, retrospective, propensity matched cohort on mortality and length of stay | Insufficient Data on Bacterial Coinfection |  |
| Cianchi, G. et al 2011 | Ventilatory and ECMO treatment of H1N1-induced severe respiratory failure: Results of an Italian referral ECMO center | <50 Influenza Cases |  |
| Cidre Lopez, R. et al 2016 | Respiratory viral infections in hematologic patients in our environment. a retrospective study | Abstract Only |  |
| Cilloniz, C. et al. 2011 | Bacterial co-infections in community acquired pneumonia cases of 2009 pandemic-influenza A (H1N1) virus in Spain | Abstract Only |  |
| Claverias, L. et al. 2019 | Effect of macrolide treatment in mortality in patients with severe pneumonia due to influenza virus infection | Abstract Only |  |
| Cordero, E. et al. 2012 | Pandemic influenza A(H1N1) virus infection in solid organ transplant recipients: impact of viral and non-viral co-infection. | Insufficient Data on Bacterial Coinfection |  |
| Crotty, MP. et al. 2015 | Epidemiology, Co-Infections, and Outcomes of Viral Pneumonia in Adults An Observational Cohort Study | Insufficient Data on Bacterial Coinfection |  |
| Dong, L. et al. 2024 | Editorial: The biological mechanism and health effect of co-infection with multiple pathogens | Abstract Only | Editorial |
| Echenique, IA. et al. 2013 | Clinical Characteristics and Outcomes in Hospitalized Patients with Respiratory Viral Co-Infection during the 2009 H1N1 Influenza Pandemic | Ineligible Outcomes |  |
| El Baroudy, NR. et al 2018 | Respiratory viruses and atypical bacteria co-infection in children with acute respiratory infection | <50 Influenza Cases |  |
| Esterman, EE. et al 2013 | Influenza infection in infants aged <6 months during the H1N1-09 pandemic: A hospital-based case series | <50 Influenza Cases |  |
| Fatima, A. et al 2019 | NH1N1 influenza and risk of bacterial co infections among acutely ill children | Abstract Only |  |
| Fawkner-Corbett, D. et al 2011 | The impact of H1N1 influenza on acute respiratory infection in pre-school children in north east Brazil | Abstract Only |  |
| Fiore-Gartland, A. et al. 2017 | Cytokine profiles of severe influenza virus-related complications in children | Insufficient Data on Bacterial Coinfection |  |
| Fu, C. et al 2025 | Clinical characteristics and co-infection analysis of influenza a virus in pediatric respiratory infections: a study based on tNGS technology | Insufficient Data on Bacterial Coinfection |  |
| Ghani, ASA. et al. 2012 | An investigation into the prevalence and outcome of patients admitted to a pediatric intensive care unit with viral respiratory tract infections in Cape Town, South Africa | Ineligible Study Design | Not all Influenza cases confirmed |
| Gu, B. et al. 2022 | Comparison of hospitalized patients with severe pneumonia caused by COVID-19 and influenza A (H7N9 and H1N1): A retrospective study from a designated hospital | <50 Influenza Cases |  |
| Gomez, L. et al 2019 | CLINICAL CHARACTERISTICS OF CRITICAL CARE PATIENTS WITH CONFIRMED DIAGNOSIS OF INFLUENZA PNEUMONIA AT THE UNIVERSITY HOSPITAL FUNDACION SANTA FE DE BOGOTA | Abstract Only |  |
| Gucchait, P. et al. 2021 | Detection of atypical pathogens in community acquired pneumonia by indirect immunofluorescence assay | <50 Influenza Cases |  |
| Holter, JC. et al 2015 | Etiology of community-acquired pneumonia and diagnostic yields of microbiological methods: a 3-year prospective study in Norway | <50 Influenza Cases |  |
| Hong, HL. Et al 2014 | Viral infection is not uncommon in adult patients with severe hospital-acquired pneumonia | <50 Influenza Cases |  |
| Hon, KL et al. 2010 | Influenza and parainfluenza associated pediatric ICU morbidity | <50 Influenza Cases |  |
| Houying, Q. et al. 2020 | Clinical characteristics and prognosis analysis of 37 patients with severe influenza | <50 Influenza Cases |  |
| Ho, Z. et al. 2015 | Clinical differences between respiratory viral and bacterial mono- and dual pathogen detected among Singapore military servicemen with febrile respiratory illness | Insufficient Data on Bacterial Coinfection |  |
| Hsu, CH. et al. 2015 | Detection of influenza and non-influenza respiratory viruses in lower respiratory tract specimens among hospitalized adult patients and analysis of the clinical outcome | <50 Influenza Cases |  |
| Huijskens, EG. et al. 2014 | The value of signs and symptoms in differentiating between bacterial, viral and mixed aetiology in patients with community-acquired pneumonia | <50 Influenza Cases |  |
| Jennings, LC. et al. 2008 | Incidence and characteristics of viral community-acquired pneumonia in adults | <50 Influenza Cases |  |
| Johansson, N. et al. 2011 | Clinical impact of combined viral and bacterial infection in patients with community-acquired pneumonia. | Insufficient Data on Bacterial Coinfection |  |
| Jung, J. et al. 2020 | Clinical significance of viral-bacterial codetection among young children with respiratory tract infections: Findings of RSV, influenza, adenoviral infections | <50 Influenza Cases |  |
| Kalasikam, M. et al. 2024 | Antibiotic Overuse in Pediatric Patients Hospitalized with RSV, COVID-19, and Influenza | Insufficient Data on Bacterial Coinfection |  |
| Kalil, AC. et al. 2019 | Influenza virus-related critical illness: Pathophysiology and epidemiology | Ineligible Study Design | Review |
| Kang, VJW. et al. 2024 | CT findings of 144 in-hospital patients with influenza pneumonia: A retrospective analysis | Insufficient Data on Bacterial Coinfection |  |
| Katzen, J. et al. 2015 | Timing of antiviral therapy in hospitalized patients with influenza is associated with clinical outcomes: A five year retrospective study | Insufficient Data on Bacterial Coinfection |  |
| Khan, ZA. et al. 2025 | Frequency of viral etiology in community-acquired pneumonia | Insufficient Data on Bacterial Coinfection |  |
| Kharosi, ZA. et al. 2017 | Epidemiological and clinical characteristics of respiratory viral infections in adults at sultan qaboos university hospital (SQUH), in Oman | Abstract Only |  |
| Kim, HJ. et al. 2018 | Respiratory virus of severe pneumonia in South Korea: Prevalence and clinical implications | <50 Influenza Cases |  |
| Kim, YN. et al. 2012 | Clinical significance of pleural effusion in the new influenza A (H1N1) viral pneumonia in children and adolescent | Insufficient Data on Bacterial Coinfection |  |
| Krittigamas, P. et al. 2020 | Differentiating Viral and Bacterial Pneumonia by Clinical Manifestations among Children in Chiang Mai, Thailand | Insufficient Data on Bacterial Coinfection |  |
| Krolikowski, K. et al. 2024 | The Proof Is in the Sputum: Untangling Viral Pneumonia From Bacterial Viral Co-infection | Abstract Only |  |
| Laundy, M. et al. 2003 | Influenza A community-acquired pneumonia in East London infants and young children. | <50 Influenza Cases |  |
| Lee, EH. et al. 2010 | Fatalities associated with the 2009 H1N1 influenza A virus in New York city | <50 Influenza Cases |  |
| Lee, N. et al. 2011 | Complications and outcomes of pandemic 2009 influenza A (H1N1) virus infection in hospitalized adults: How do they differ from those in seasonal influenza? | Insufficient Data on Bacterial Coinfection | Only propensity scores, no raw data. |
| Lee, N. et al. 2012 | Diagnosis, management and outcomes of adults hospitalized with influenza | No Access |  |
| Leli, C. et al. 2021 | Prevalence of respiratory viruses by multiplex pcr: A four-and-a-half year retrospective study in an italian general hospital | Insufficient Data on Bacterial Coinfection |  |
| Levy, E. et al. 2018 | Association of mannose-binding lectin with influenza critical illness in children | Abstract Only |  |
| Li, M. et al. 2023 | Metagenomic-based pathogen surveillance for children with severe pneumonia in pediatric intensive care unit | <50 Influenza Cases |  |
| Lin, C. et al. 2019 | Etiology and characteristics of community-acquired pneumonia in an influenza epidemic period | <50 Influenza Cases |  |
| Liu, J. et al. 2015 | Prevalence and correlation of infectious agents in hospitalized children with acute respiratory tract infections in central China | Insufficient Data on Bacterial Coinfection |  |
| Liu, WJ. et al. 2018 | Clinical, immunological and bacteriological characteristics of H7N9 patients nosocomially co-infected by Acinetobacter Baumannii: a case control study | Insufficient Data on Bacterial Coinfection |  |
| Li, XD. et al. 2023 | Epidemiological investigation of lower respiratory tract infections during influenza A (H1N1) pdm09 virus pandemic based on targeted next-generation sequencing | <50 Influenza Cases |  |
| Loevinsohn, G. et al. 2021 | Respiratory pathogen diversity and co-infections in rural Zambia | Insufficient Data on Bacterial Coinfection |  |
| Lv, G. et al. 2024 | Epidemiological characteristics of common respiratory pathogens in children | Insufficient Data on Bacterial Coinfection |  |
| MacIntyre, CR. et al. 2017 | Viral and bacterial upper respiratory tract infection in hospital health care workers over time and association with symptoms | Insufficient Data on Bacterial Coinfection |  |
| Malato, L. et al. 2011 | Pandemic influenza A(H1N1) 2009: molecular characterisation and duration of viral shedding in intensive care patients in Bordeaux, south-west France, May 2009 to January 2010 | <50 Influenza Cases |  |
| Martinez, A. et al. 2021 | Antibiotic Prescribing Trends in Hospitalized Influenza Versus COVID-19 Patients at a Community-Based Health System | Abstract Only | Poster Abstract |
| Martin-Loeches, I. et al. 2013 | Macrolide-based regimens in absence of bacterial co-infection in critically ill H1N1 patients with primary viral pneumonia | Insufficient Data on Bacterial Coinfection |  |
| Martin-Loeches, I. et al. 2011 | Community-acquired respiratory coinfection in critically ill patients with pandemic 2009 influenza A(H1N1) virus | No Access |  |
| May, A. et al. 2021 | Bacterial and fungal respiratory co-infection among patients admitted to ICU with COVID-19: A retrospective cohort study in a UK hospital | Ineligible Study Design | Only SARS-CoV-2 |
| McFarlane, A. et al. 2015 | Hospitalized influenza patients during 2013-2014: A comparison of ICU and ward treated patients including antimicrobial therapy, adverse events, and outcomes | No Access | No DOI |
| Miggins, M. et al. 2011 | The potential influence of common viral infections diagnosed during hospitalization among critically ill patients in the United States | Insufficient Data on Bacterial Coinfection |  |
| Mikkelsen, VS. et al. 2021 | COVID-19 versus influenza A/B supeRInfectionS in the IntenSive care unit (CRISIS): Protocol for a Danish nationwide cohort study | Ineligible Study Design | Protocol |
| Miroballi, Y. et al. 2010 | Novel influenza A(H1N1) in a Pediatric Health Care Facility in New York City during the first wave of the 2009 pandemic | Insufficient Data on Bacterial Coinfection |  |
| Miron, VD. et al. 2016 | Pneumococcal superinfection in children with influenza | Abstract Only |  |
| Mombelli, M. et al. 2019 | Burden of respiratory virus infections in solid-organ transplant recipients: A nationwide multi-season cohort study | Abstract Only |  |
| Moolasart, V. et al. 2024 | Prevalence of Co-Infections and Pathogens in Hospitalized Children with Acute Respiratory Infections: A Comparative Analysis Between SARS-CoV-2 and Non-SARS-CoV-2 Cases | <50 Influenza Cases |  |
| Musher, DM. et al. 2021 | Bacterial coinfection in COVID-19 and influenza pneumonia | Abstract Only | Letter to the Editor |
| Naderi, H. et al. 2015 | Etiological diagnosis of community-acquired pneumonia in adult patients: A prospective hospital-based study in Mashhad, Iran | <50 Influenza Cases |  |
| Nguyen Van Tam, JS. et al. 2010 | Risk factors for hospitalisation and poor outcome with pandemic A/H1N1 influenza: United Kingdom first wave (May-September 2009) | Insufficient Data on Bacterial Coinfection | Relies on Radiography, incomplete Microbiology data. |
| Odongo, FCA. et al 2016 | Clinical Characteristics and Outcomes of Influenza A Infection in Kidney Transplant Recipients: A Single-Center Experience | <50 Influenza Cases |  |
| O’Leary, ST. et al. 2024 | Recommendations for Prevention and Control of Influenza in Children, 2024-2025: Technical Report | Insufficient Data on Bacterial Coinfection |  |
| O’Neal, J. et al. 2021 | Incidence of community acquired pneumonia with viral infection in mechanically ventilated patients in the medical intensive care unit | Abstract Only |  |
| Ostby, AC. et al. 2012 | The clinical significance of respiratory viruses and coinfections in hematological patients | No Access | No DOI |
| Østby, AC. et al. 2013 | Respiratory virology and microbiology in intensive care units: A prospective cohort study | <50 Influenza Cases |  |
| Oztelcan Gunduz, B. et al. 2021 | Evaluation of Influenza Patients Admitted in 2019-2020 Flu Season | Ineligible Patient Population |  |
| Oliva, A. et al. 2021 | Comparison of clinical features and outcomes in COVID-19 and influenza pneumonia patients requiring intensive care unit admission | <50 Influenza Cases |  |
| Pandey, M. et al. 2022 | Comparative incidence of early and late bloodstream and respiratory tract co-infection in patients admitted to ICU with COVID-19 pneumonia versus Influenza A or B pneumonia versus no viral pneumonia: wales multicentre ICU cohort study | <50 Influenza Cases |  |
| Patel, N. et al. 2022 | Epidemiology and Outcomes of Bacterial Coinfection in Hospitalized Children With Respiratory Viral Infections: A Single Center Retrospective Chart Review | Ineligible Outcomes |  |
| Perez-Garcia, F. et al. 2016 | Influenza A and B co-infection: a case–control study and review of the literature | Insufficient Data on Bacterial Coinfection |  |
| Pugachev, M. et al. 2015 | The role of viral infection in the progression of community-acquired pneumonia (CAP) | Abstract Only |  |
| Qu, J. et al. 2022 | Aetiology of severe community acquired pneumonia in adults identified by combined detection methods: a multi-centre prospective study in China | Ineligible Outcomes |  |
| Quah, J. et al. 2018 | Impact of microbial Aetiology on mortality in severe community-acquired pneumonia | Ineligible Outcomes |  |
| Randolph, AG. et al. 2010 | Risk factors for mortality in children admitted to the ICU with influenza infection | Abstract Only |  |
| Reed, C. et al. 2009 | Infection with community-onset Staphylococcus aureus and influenza virus in hospitalized children | Insufficient Data on Bacterial Coinfection |  |
| Reyes, LF. et al. 2020 | Bacterial co-infection in patients admitted to the ICU due to severe influenza is not related with previous influenza vaccination | Abstract Only |  |
| Rice, TW. et al. 2011 | Role of bacterial co-infection on illness severity and outcomes in ICU patients with 2009 pandemic influenza a (H1N1) infection | Abstract Only |  |
| Rodriguez, A. et al. 2024 | A Machine Learning Approach to Determine Risk Factors for Respiratory Bacterial/Fungal Coinfection in Critically Ill Patients with Influenza and SARS-CoV-2 Infection: A Spanish Perspective | Re-analysis |  |
| Rosculet, C. et al. 2016 | The severity of A H1N1 Influenza infection in the 2015-2016 season | Abstract Only |  |
| Rozencwajg, S. et al. 2017 | Bacterial co-infection in influenza-associated ARDS supported by ECMO | Abstract Only |  |
| Ryan, D. et al. 2014 | Aetiology of community-acquired pneumonia in the ICU setting and its effect on mortality, length of mechanical ventilation and length of ICU stay: A 1-year retrospective review | <50 Influenza Cases |  |
| Rzymski, P. et al. 2025 | Unraveling Poland's unprecedented influenza surge in early 2025: increased viral severity or post-pandemic vulnerability? | Insufficient Data on Bacterial Coinfection |  |
| Saez Garcia, L. et al. 2022 | INFLUENZA VIRUS INFECTION IN CHILDREN ADMITTED TO THE PEDIATRIC INTENSIVE CARE UNIT | <50 Influenza Cases |  |
| San Gil, A. et al. 2011 | Aetiology of community-acquired pneumonia in the adult and the H1N1 influenza pandemic | Abstract Only |  |
| Schrag, SJ. et al. 2006 | Multistate surveillance for laboratory-confirmed, influenza-associated hospitalizations in children 2003-2004 | Insufficient Data on Bacterial Coinfection |  |
| Scott, J. et al. 2016 | 2015/16 influenza pneumonitis: Mater misericordiae university hospital intensive care unit | Abstract Only |  |
| Shalabi, RD. et al. 2022 | Predictors of unfavorable outcome in children hospitalized with influenza and differences in clinical presentation among serotypes | Insufficient Data on Bacterial Coinfection |  |
| Shannon, KL. et al. 2022 | Viral co-infections are associated with increased rates of hospitalization in those with influenza | Insufficient Data on Bacterial Coinfection |  |
| Shen, XY. et al. 2022 | Concomitant viral and bacterial pneumonia among patients in ICU with mechanical respiratory support | <50 Influenza Cases |  |
| Shibli, F. et al. 2010 | Etiology of community-acquired pneumonia in hospitalized patients in northern Israel | <50 Influenza Cases |  |
| Sietses, M. et al. 2013 | High incidence of respiratory viruses in critically ill adult patients with respiratory failure | Abstract Only |  |
| Solis-Esquilin, EA. et al. 2020 | Profile of Hispanic Patients admitted to a Pediatric Intensive Care Unit in a community hospital due to influenza | Abstract Only |  |
| Song, JY. et al. 2011 | Clinical, laboratory and radiologic characteristics of 2009 pandemic influenza A/H1N1 pneumonia: Primary influenza pneumonia versus concomitant/secondary bacterial pneumonia | Insufficient Data on Bacterial Coinfection |  |
| Sun, RY. et al. 2024 | Influenza A/H3N2 and Its Co-infection with Other Respiratory Pathogens: Higher Pneumonia Rates and Prolonged Hospital Stays inPediatric Patients | Insufficient Data on Bacterial Coinfection |  |
| Taylor, C. et al. 2011 | H1N1: The co-infection conundrum | No Access | No DOI |
| Tief, F. et al. 2016 | An inception cohort study assessing the role of pneumococcal and other bacterial pathogens in children with influenza and ILI and a clinical decision model for stringent antibiotic use | Ineligible Study Design | Unclear distinction between ILI and confirmed Influenza |
| Tsai, YW. et al. 2023 | The risk of methicillin-resistant Staphylococcus aureus infection following COVID-19 and influenza: A retrospective cohort study from the TriNetX network | Abstract Only | Letter to the editor |
| Vaz, JA. et al. 2019 | WHEN INFLUENZA INFECTION COMPLICATES - ANALYSIS OF HOSPITALISATIONS IN INTERNAL MEDICINE DEPARTMENT | Ineligible Study Design |  |
| Vitalpur, G. et al. 2013 | Innate immune function and mortality in critically ill children with influenza: A multicenter study | Abstract Only |  |
| Voiriot, G. et al. 2016 | In severe pneumonia, respiratory viruses are not passengers but pathogens | Abstract Only |  |
| Wang, XJ. et al. 2010 | [The analysis of the clinical features between survivors and non-survivors with the severe form of new influenza A (H1N1) viral infection]. | No Access | No DOI |
| Weidmann, MD. et al. 2022 | Prevalence and Clinical Disease Severity of Respiratory Coinfections During the Coronavirus Disease 2019 Pandemic | Insufficient Data on Bacterial Coinfection |  |
| Williams, DJ. et al. 2011 | Influenza Coinfection and Outcomes in Children With Complicated Pneumonia | <50 Influenza Cases |  |
| Windsor, WJ. et al. 2022 | Clinical characteristics and illness course based on pathogen among children with respiratory illness presenting to an emergency department | Insufficient Data on Bacterial Coinfection |  |
| Wuister, AMH. et al. 2017 | Characteristics of critically ill influenza patients in an intensive care unit in the Netherlands | <50 Influenza Cases |  |
| Xu, YS. et al. 2025 | Factors affecting the severity of respiratory infections: a hospital-based cross-sectional study | Insufficient Data on Bacterial Coinfection |  |
| Yan, XX. et al. 2011 | Influenza a (H1n1) virus complicated with bacteria infection | No Access | No DOI |
| Yao, Y. et al. 2019 | Epidemiology of pathogens causing acute respiratory infections in children in Beijing during 2016 to 2018 | No Access |  |
| Yildirim, M. et al. 2020 | Comparison of critically-Ill COVID-19 and influenza patients with acute respiratory failure | Abstract Only |  |
| Yoo, JW. et al. 2019 | Characteristics and outcomes of patients with pulmonary acute respiratory distress syndrome infected with influenza versus other respiratory viruses | <50 Influenza Cases |  |
| Zakharenkov, IA. et al. 2020 | Etiology of severe community-acquired pneumonia in adults: Results of the first Russian multicenter study | Non-English Full Text | Russian full text |

| **Supplementary Table 21 – Funding Information of included studies.**  Where the data was available it was copied, unedited, from each paper or publisher website. | | |
| --- | --- | --- |
| **Study** | **Funding Information** | **Conflict of Interest** |
| Abelenda-Alonso G, et al., 2020 | Funded by La Marato de TV3 project 201808-10; institutional support from CERCA Programme/Generalitat de Catalunya. | G.A.A. reported grants from TV3-Fundacion La Marato outside the submitted work; relevant conflicts disclosed. |
| Ahn S, et al., 2011 | N/A | N/A |
| Al Ali A, et al. 2021 | No specific grant from public or commercial funding agencies. | No conflict of interest declared. |
| Al-Dorzi H.M, et al. 2024 | No direct or indirect financial contribution/support from any organization/donor. | No competing interests declared. |
| Anania V.G, et al. 2020 | This work was supported by the National Institutes of Health (R01AI084011 and R21HD095228, A.R.), the Centers for Disease Control and Prevention (A.R.), and Genentech, Inc. | The authors declare that this study received funding from Genentech. V.A., A.N., X.Y., J.M., W.R.M., and C.R. are employees of Genentech. The funder had no role in study design, data collection and analysis, decision to publish, or preparation of the manuscript. A.R. has received research funding from Genentech, Inc., to her institution. All authors have submitted the ICMJE Form for Disclosure of Potential Conflicts of Interest. Conflicts that the editors consider relevant to the content of the manuscript have been disclosed. |
| Bartley P.S, et al. 2022 | The study was supported by the Agency for Health Research and Quality (AHRQ grant no. 5 R01 HS024277-05) to M.B.R. | M.K. has received grant funding from the Centers for Disease Control and Prevention (CDC) and royalties from UpToDate for chapters on hospital-acquired pneumonia. A.D. has received research support from the AHRQ, research support from The Clorox Company; he serves as a consultant to Merck. M.Z. has received research support from Tetraphase Pharmaceuticals, Astellas, Lungpacer, Merck, Spero, The Medicines Company, and Melinta; she has also received consulting fees from Paratek, Arasanis, Shionogi, Pfizer, Nabriva, and Melinta. All other authors report no conflicts of interest relevant to this article. |
| Bender J.M, et al. 2010 | J.M.B. is the recipient of a NIH Rocky Mountain Regional Center for Excellence in Biodefense and Emerging Diseases young investigator award U54 AI065357. P.G. and C.L.B. are supported in part through the CDC-funded Center of Excellence in Public Health Informatics (CDC 1 PO1 CD000284) at the University of Utah. R.S. is the recipient of a NIH/Eunice Kennedy Shriver NICHD career development award K23 HD052553. C.L.B. is further supported by grants from the Public Health Services research grant UL1-RR025764 from the National Center for Research Resources (NIH/NIAID 1 U01 AI074419 and U01-A1061611), and the NIH/Eunice Kennedy Shriver NICHD K24- HD047249. This project was in addition supported by the Children's Health Research Center at the University of Utah. | N/A |
| Beumer M.C, et al. 2019 | Funding not applicable. | No competing interests declared. |
| Bjarnason A, et al. 2012 | This work was supported by grants from the Icelandic Center for Research, Rannís [grant number 100436021]; and the Landspitali University Hospital Science Fund. URL: http://rannis.is/english/home/. The funders had no role in study design, data collection and analysis, decision to publish, or preparation of the manuscript. | The authors have declared that no competing interests exist. |
| Blyth C.C, et al. 2013 | Funding for the study was provided from a untied grant through the investigator-initiated grants program of Merck US – grant #IISP3732. | N/A |
| Carbonell R, et al. 2021 | This research received no external funding. This study was supported by the Spanish Intensive Care Society (SEMICYUC) and Ricardo Barri Casanovas Foundation (Alejandro Rodríguez). The study sponsors have no role in the study design, data collection, data analysis, data interpretation, or writing of the report. | Patient consent was waived due to the anonymization of all collected data. |
| Chen J, et al. 2018 | This study was supported by National Mega Projects of Science and Technology in 13th Five-Year Plan of China: Technical Platform for Communicable Disease Surveillance Project (2017ZX10103010–002). | The authors declare that they have no competing interests. |
| Choi S.H, et al. 2015 | This study was supported by grant 2012-389 from the Asan Institute of Life Sciences, Seoul, Republic of Korea. | The authors declare that there are no conflicts of interest to disclose. |
| Christensen I, et al. 2023 | This research received no external funding, but was funded internally from the Østfold Hospital Thrust. | The authors declare no conflict of interest. The funders had no role in the design of the study; in the collection, analyses, or interpretation of data; in the writing of the manuscript; or in the decision to publish the results. |
| Cillóniz C, et al. 2012 | Supported by CibeRes (CB06/06/0028), 2009 SGR 911, Programa de Investigacion en Gripe-ISCiii-McyT, and IDIBAPS. | N/A |
| Cordero E, et al. 2012 | Supported by Ministerio de Ciencia e Innovacion/Instituto de Salud Carlos III Programa de Investigacion Sobre Gripe A/H1N1 (GR09/0014), REIPI RD06/0008 co-financed by ERDF, and IDIBELL grant to D. Viasus. | N/A |
| Cuquemelle E, et al. 2011 | The REVA-SRLF registry was supported by grants from the Société de Réanimation de Langue Française (SRLF), the French Research Agency (ANRS) and the French Ministry of Health. | N/A |
| D'Onofrio V, et al. 2021 | FAPIC project funded by EU Horizon 2020 research and innovation program, GA 634137. | No conflict of interest declared. |
| Damasio G.A, et al. 2015 | N/A | declare have no competing ﬁnancial interests. |
| Dawood F.S, et al. 2010 | N/A | The authors declare no conflicts of interest. |
| Dhanoa A, et al. 2011 | Supported by grants from Karolinska Institutet. | No conflict of interests relevant to the paper reported. |
| Dumas G, et al. 2023 | Funding: none. | G.D. received grants from HoldEm For Life Oncology Award and Societe de Reanimation en Langue Francaise. |
| Eşki A, et al. 2019 | No funding disclosed; authors state no funding to disclose. | No conflicts of interest to disclose. |
| Estenssoro E, et al. 2010 | Supported by the Argentinian Society of Intensive Care Medicine. | N/A |
| Falsey A.R, et al. 2013 | This work was supported by the National Institute of Allergy and Infectious Diseases (1R01AI079446-01). | Dr Falsey has received research grants from Medimmune/AstraZeneca and sanofipasteur and consulting fees from Medimmune/AstraZeneca, sanofipasteur, GlaxoSmithKline, and Novavax. Dr Walsh has received consulting fees from Medimmune/AstraZeneca, GlaxoSmithKline, Novartis, Microdose, Alios Pharmaceuticals, Alnylam, and ClearPath. All other authors report no potential conflicts. |
| Feys S, et al. 2024 | Funding acquisition noted for S.H.-B., G.V.V., J.V.W., T.G., J.W., and A.C.; specific funders not detailed in extracted PDF text. | Author disclosures available with article at ATS website; detailed disclosures not in PDF text. |
| Garg S, et al. 2015 | N/A | The authors declare that they have no competing interests. |
| Guo L, et al. 2019 | This work was supported by National Key R&D Program of China (Grant Nos. 2017YFC1309700 and 2017YFC1309701), by the National Natural Science Foundation of China (Grant No. 81570029), and by Shanghai Key Discipline for Respiratory Diseases (Grant No. 2017ZZ02014). This work was also funded in part by a grant from Innovative research team of high-level local universities in Shanghai, and by Institute of Respiratory Disease, School of Medicine, Shanghai Jiao Tong University. | The authors declare that the research was conducted in the absence of any commercial or financial relationships that could be construed as a potential conflict of interest. |
| Gutiérrez-Pizarraya A, et al. 2012 | This work was supported by the Ministerio de Ciencia e Innovación, Instituto de Salud Carlos III (GR09/0041) and - co-financed by European Development Regional Fund "A way to achieve Europe" ERDF, Spanish Network for the Research in Infectious Diseases (REIPI RD06/0008). P.P-R. was funded by Instituto de Salud Carlos III, Programme Miguel Servet (CP05/00226). | N/A |
| Haddara A, et al. 2024 | No specific grant from public, commercial, or not-for-profit funding agencies. | No established conflicting financial interests or personal relationships declared. |
| Haeberer M, et al. 2024 | This study was funded by Pfizer Ltd, including the journal's Rapid Service Fee. | Mariana Haeberer, Rong Fan, Qing Liu, Sonal Uppal, Caihua Liang, and Elizabeth Begier are employees of Pfizer and may own Pfizer stock. Yongzheng He has been hired by Pfizer to provide support for statistical analysis. Marina Toquero Asensio, Alejandro Martín Toribio, Silvia Rojo-Rello, Marta Domínguez-Gil, Cristina Hernán-García, Virginia Fernández-Espinilla, José M. Eiros, Javier Castrodeza Sanz, and Ivan Sanz-Muñoz received funding from Pfizer for data abstraction and manuscript review. |
| Hagerman A, et al. 2015 | No financial support reported. | No potential conflict of interest reported. |
| Hall M.W, et al. 2013 | Supported, in part, by the Centers for Disease Control and Prevention, Atlanta, GA; America’s Health Insurance Plans Foundation; the Health Respiratory Research Network of the Fonds de Recherche en Santé du Québec; the National Institutes of Health (AI084011); and the Research Institute at Nationwide Children’s Hospital. This work represents the findings of the authors and not necessarily the views of the Centers for Disease Control and Prevention or the National Institutes of Health. | The authors have not disclosed any potential conflicts of interest. |
| Hayashi Y, et al. 2012 | None. | Dr David L. Paterson has been on advisory boards for Merck, AstraZeneca, Cubist, Pfizer and Leo Pharmaceuticals. Dr Graeme R. Nimmo has been on advisory boards on Pfizer and Wyeth. None of the other authors has any conflict of interest. |
| Hedberg P, et al. 2022 | N/A | N/A |
| Hernandez-Bou S, et al. 2013 | N/A | The authors declare no conflict of interest. |
| Hsing T, et al. 2022 | This research was supported in part by the Ministry of Science and Technology, Taiwan with grant nos. MOST 109-2314-B-002-238 to LY Chang. | The authors have no conflicts of interest relevant to this article. |
| Ishiguro T, et al. 2013 | N/A | The authors state that they have no Conflict of Interest (COI). |
| Jamoussi A, et al. 2022 | No specific funding received. | No competing interests declared. |
| Jorda A, et al. 2024 | Open access funding provided by Medical University of Vienna; research funded by Medical Scientific Fund of the Mayor of the City of Vienna (Project ID 22153). | No competing interests declared. |
| Khandaker G, et al. 2014 | This project was funded by the National Health and Medical Research Council (NHMRC) of Australia (H1N1 Grant no: 633028) and the NSW Department of Health. Activities of the NCIRS are supported by the Australian Government Department of Health and Ageing (DoHA). APSU is supported by the DoHA; the NHMRC (Enabling Grant No: 402784 and Practitioner Fellowship No: 457084, E. Elliott); the Creswick Foundation (Fellowship: Y. Zurynski); the Discipline of Paediatrics and Child Health and the Sydney Medical School of the University of Sydney; the Children's Hospital at Westmead; and the Royal Australasian College of Physicians. G. Khandaker was supported by an NHMRC Project Grant (No. 633032). H. Marshall is supported by an NHMRC Career Development Fellowship (No. 1016272). | GK was an investigator in studies supported by Roche. HM has participated as a Board Member of Global Advisory Boards for Merck, Novartis and GlaxoSmithKline, has received travel support to present scientific data at international meetings and is an investigator in industry-sponsored clinical vaccine trials. Her institution has received funding for investigator-led research from Novartis, Pfizer and GlaxoSmithKline. JB has served on advisory and/or data safety monitoring committees for CSL Vaccines and GSK for which the Murdoch Children's Research Institute (MCRI) receives payments into an educational fund. MCRI has received travel support for JB to present scientific data at international meetings, and he is an investigator in industry-sponsored vaccine trials. PCR is a member of the Vaccine Trials Group. The Vaccine Trials Group has received institutional funding for research from vaccine providers including CSL Biotherapies, Sanofi Pasteur, Baxter and GlaxoSmithKline. PCR has also previously been a member of a CSL Limited vaccine scientific advisory board and received travel support from Baxter, Sanofi and Pfizer to present scientific data at international meetings. RB has received financial support from CSL, Sanofi, GSK, Roche, Novartis and Wyeth, to conduct research and present at scientific meetings. Any funding received is directed to an NCIRS research account at The Children's Hospital at Westmead and is not personally accepted by Professor Booy. The other authors declare that they have no conflict of interests in relation to this work. |
| Kim H.S, et al. 2011 | N/A | We declare that we have no conflicts of interest and financial support. We are grateful to the EIS officers in Korea who supplied a standardized case report of deaths associated with 2009 pandemic influenza A, Mina Baek for managing the database, Hochun Choi and Jinsu Song for writing assistance, and members in The Division of KCDC's Infectious Disease Surveillance for providing estimated case numbers. |
| Kim J.H, et al. 2018 | N/A | The authors have no conflicts of interest to declare. |
| Kim S.H, et al. 2011 | None of the authors has a financial relationship with a commercial entity that has an interest in the subject of this manuscript. | None of the authors has a financial relationship with a commercial entity that has an interest in the subject of this manuscript. |
| Krammer M, et al. 2024 | No external funding was received for this investigation. | The authors declare no competing interests. |
| Kumar S, et al. 2010 | N/A | N/A |
| Kwon Y.S, et al. 2017 | Supported by National Research Foundation of Korea grant funded by Korean Government (MSIP), No. 2014R1A5A2010008. | No competing interests declared. |
| Le Glass E, et al. 2022 | This work was supported by the French Government under the ‘Investments for the Future’ programme managed by the National Agency for Research (ANR), Méditerranée-Infection 10-IAHU-03 and was also supported by Région Provence Alpes Côte d'Azur and European FEDER PRIMMI funding (European Regional Development Fund–Plateformes de Recherche et d'Innovation Mutualisées Méditerranée Infection), ERDF PA 0000320 PRIMMI. | Didier Raoult has been a consultant for Hitachi High-Technologies Corporation, Tokyo, Japan, from 2018 to 2020. He is a scientific board member of Eurofins company and a founder of a microbial culture company (Culture Top). Other authors declare no conflicts of interest. |
| Lee W.C, et al. 2022 | None. | The authors declare that they have no competing interests. |
| Li Y.N, et al. 2025 | All phases of this study were supported by Beijing Research Center for Respiratory Infectious Diseases Project (BJRID2025-008) and 2022 Key Specialty Construction Project for Prevention and Control of Major Epidemics in Beijing, China (2−1−2−6−15). | The authors declare that they have no known competing financial interests or personal relationships that could have appeared to influence the work reported in this paper. |
| Li Z, et al. 2019 | N/A | N/A |
| Liderot K, et al. 2013 | Supported by grants from Karolinska Institutet. | No conflict of interests relevant to the paper reported. |
| Liu J, et al. 2020 | N/A | Conflicts of interest: All the authors report no conflict of interest relevant to this article. |
| Liu Y, et al. 2021 | Commissioned Programmes for Influenza Research, Health and Medical Research Fund (HMRF), FHB (Ref. No.: INF-CUHK-2);National Natural Science Foundation of China (81873560);Shenzhen Science and Technology Programme;Health and Medical Research Fund (18170092). | Lowell Ling has received consulting fees from Merck Sharp & Dohme. Other authors declared that they have no conflict of interest. |
| Lopez-Delgado J.C, et al. 2013 | N/A | N/A |
| Lu Y, et al. 2013 | This study was supported by Grants from the State Major Infectious Disease Research Program (China Central Government, 2009ZX10004206 and 2012ZX10004206) and the Key Projects of Nature Science Fund of Shandong Province (ZR2009CZ011). | The authors declare that there is no conflict of interests regarding the publication of this paper. |
| Mangas-Moro A, et al. 2023 | This research did not receive any specific grants from public sector agencies, commercial entities, or non-profit organizations. | None. |
| Marcoux D, et al. 2022 | This research received no external funding. | The authors declare no conflict of interest. |
| Marin-Corral J, et al. 2018 | The registry of patients with influenza A was developed by the GETGAG and is the property of the SEMICYUC. | The authors have no conflict of interest to declare. |
| Martin-Loeches I, et al 2017 | The study funder (Spanish Society of Critical Care—SEMICYUC) had no role in the study design, data collection, data analysis, data interpretation, or writing of the report. The corresponding author (IML) had full access to all the data in the study and final responsibility for the decision to submit for publication. | All of the authors declare that no conflict of interest exists. |
| Melamed K.H, et al. 2020 | No financial support received. | No potential conflicts of interest declared. |
| Mendoza M.A, et al. 2023 | No specific funding received. | No conflict of interest declared. |
| Mercat A, et al. 2011 | N/A | The authors declare that they have no conflict of interest. |
| Merișescu M.M, et al. 2023 | This research received no external funding. | The authors declare no conflict of interest. |
| Muscedere J, et al. 2013 | No funding received for the study. | N/A |
| Nguyen T, et al. 2012 | N/A | N/A |
| Nolan V.G, et al. 2018 | This work was supported by the Centers for Disease Control and Prevention (grant U18IP000489 to J. A. M.). | All authors: No reported conflicts of interest. All authors have submitted the ICMJE Form for Disclosure of Potential Conflicts of Interest. Conflicts that the editors consider relevant to the content of the manuscript have been disclosed. |
| Owayed A.F, et al. 2012 | N/A | N/A |
| Poulakou G, et al. 2011 | N/A | N/A |
| Qin T, et al. 2020 | This work was supported by the National Natural Science Foundation of China (grant number 81671985); the National Science and Technology Major Project of China (grant number 2018ZX10712001-007); the Sanming Project of Medicine in Shenzhen (grant number SZSM201811071); and the Research Units of Discovery of Unknown Bacteria and Function, Chinese Academy of Medical Sciences (grant number 2018RU010). | No potential conflict of interest was reported by the author(s). |
| Randolph A.G, et al. 2011 | The authors have indicated they have no financial relationships relevant to this article to disclose | The authors have indicated they have no financial relationships relevant to this article to disclose |
| Rice T.W, et al. 2012 | N/A | N/A |
| Rouzé A, et al. 2021 | Supported in part by French Programme Investissement d Avenir (I-SITE ULNE) via ANR coVAPid project; I.M.-L. supported by Science Foundation Ireland grant 20/COV/0038; funders had no role. | Author disclosures available with article at ATS website; detailed disclosures not in PDF text. |
| Rozencwajg S, et al. 2018 | Funding: none. | C.-E.L. reported personal fees/nonfinancial support; A.C. trial support; N.B., G.L., and A.C. honoraria from MAQUET; others no competing interests. |
| Schoettler J.J, et al. 2023 | This research was funded by the Klaus Tschira Foundation (grant number: 00.277.2015) | The authors declare no conflict of interest. |
| Shafran N, et al. 2021 | Supported by the Milner Foundation. | No competing interests declared. |
| Shah NS, et al. 2016 | Supported in part by HRSA grant D33HP25768. | Competing interests: none. |
| Shi T, et al. 2019 | The authors have no funding and conflicts of interest to disclose. | The authors have no funding and conflicts of interest to disclose. |
| Sohn C.H, et al. 2013 | Authors reported no relevant financial information. | No potential conflicts of interest reported. |
| Soler-Font M, et al. 2022 | This study was supported by the Programme of Prevention, Surveillance and Control of Transmissible Diseases (PREVICET), CIBER de Epidemiología y Salud Pública (CIBERESP, CB06/02/0076, CB16/02/00322 and CB16/02/00429), Instituto de Salud Carlos III, Madrid; and the Catalan Agency for the Management of Grants for University Research (AGAUR Grant Number 2017/SGR 1342). | The authors declare no conflict of interest. |
| Tao RJ, et al. 2018 | N/A | N/A |
| Tasar S, et al. 2022 | N/A | No conflicts of interest declared. |
| Teng F, et al. 2019 | N/A | No competing interests declared. |
| Thelen J.M, et al. 2021 | No endorsement or funding received. | No conflict of interest declared. |
| Tian J, et al. 2023 | This study was supported by grants from Guizhou Provincial Respiratory Critical Disease Clinical Research and Prevention and Treatment Talent Base Project ([2020]8), Zunyi Respiratory Medicine Talent Base Project ([2019]69), Science and Technology Fund of Guizhou Provincial Health Commission ([gzwkj2023-017]), Science and Technology Bureau Project of Zunyi City (Zunshi Kehe HZ [2020]292). The funders had no role in the study design, data collection and analysis, decision to publish, or reparation of the work. | None. |
| Tokuhira N, et al. 2012 | N/A | The authors have not disclosed any potential conflicts of interest. |
| Tsai C.F, et al. 2023 | Supported by Ministry of Science and Technology (MOST 109-2314-B-002-238, 111-2314-B-002-263) and Chi Mei Medical Center (CMOR11202, CMOR11101, CMNCKU11101, CCFHR11102); funders had no role. | No conflict of interest declared. |
| Üzüm Ö, et al. 2022 | Permission for this study was granted by the Ethics Committee with the decision numbere 2019/13-20 and dated 11 September 2019. | N/A |
| Verdier V, et al. 2023 | No funding received for the study. | No conflict of interest declared. |
| Viasus D, et al. 2011 | Supported by Ministerio de Ciencia e Innovacion/Instituto de Salud Carlos III Programa de Investigacion sobre gripe A/H1N1 (GR09/0014), REIPI RD06/0008 co-financed by ERDF, and IDIBELL grant to D. Viasus. | No dual/conflicting interest declared. |
| Viasus D, et al. 2013 | This research was supported by the Ministerio de Ciencia e Innovación, Instituto de Salud Carlos III, Programa de Investigación sobre gripe A/H1N1 (GR09/0014). It was co-financed by Ministerio de Economía y Competitividad, Instituto de Salud Carlos III – co-financed by European Development Regional Fund "A way to achieve Europe" ERDF, Spanish Network for the Research in Infectious Diseases (REIPI RD12/0015). DV is the recipient of a research grant from the REIPI. | All authors declare that they have no conflicting interests that are relevant to this article. |
| von Baum H, et al. 2011 | This network is supported by the German Ministry of Education and Research (Bundesministerium für Bildung und Forschung) Berlin, Germany. | None declared. |
| Wallemacq S, et al. 2022 | This research did not receive any specific grant from funding agencies in the public, commercial, or not-for-profit sectors. | None. |
| Wei L, et al. 2015 | This study was supported by grants from the China Mega-Project on Infectious Disease Prevention (No. 2013ZX10004202-002) and National Science Fund for Young Scholars (No. 81222037). BJC received research funding from MedImmune Inc. and Sanofi Pasteur. | BJC consults for Crucell NV. The other authors have no conflicts of interest to disclose. |
| Zhang Q, et al. 2011 | The research study and the funding for the project were approved by Lanzhou University. The study was performed in compliance with the guidelines of our Institutional Review Board of the Second Hospital of Lanzhou University. The study was also approved by Department of Pediatrics of the Second Hospital of Lanzhou University. Written informed consent was obtained from all subject's parents before the study was performed. | N/A |
| Zhang Y, et al. 2020 | Supported by National Science Fund for Distinguished Young Scholars (81425001/H0104 to Bin Cao), CAMS Innovation Fund for Medical Sciences (2018-I2M-1-003 to Bin Cao), and Intramural Research Program of China-Japan Friendship Hospital (2018-2-QN-22 to Yulin Zhang). | No conflict of interest declared. |
| Zhong PP, et al. 2016 | Zhejiang Provincial Department of Science and Technology Project (2015C37026) | N/A |

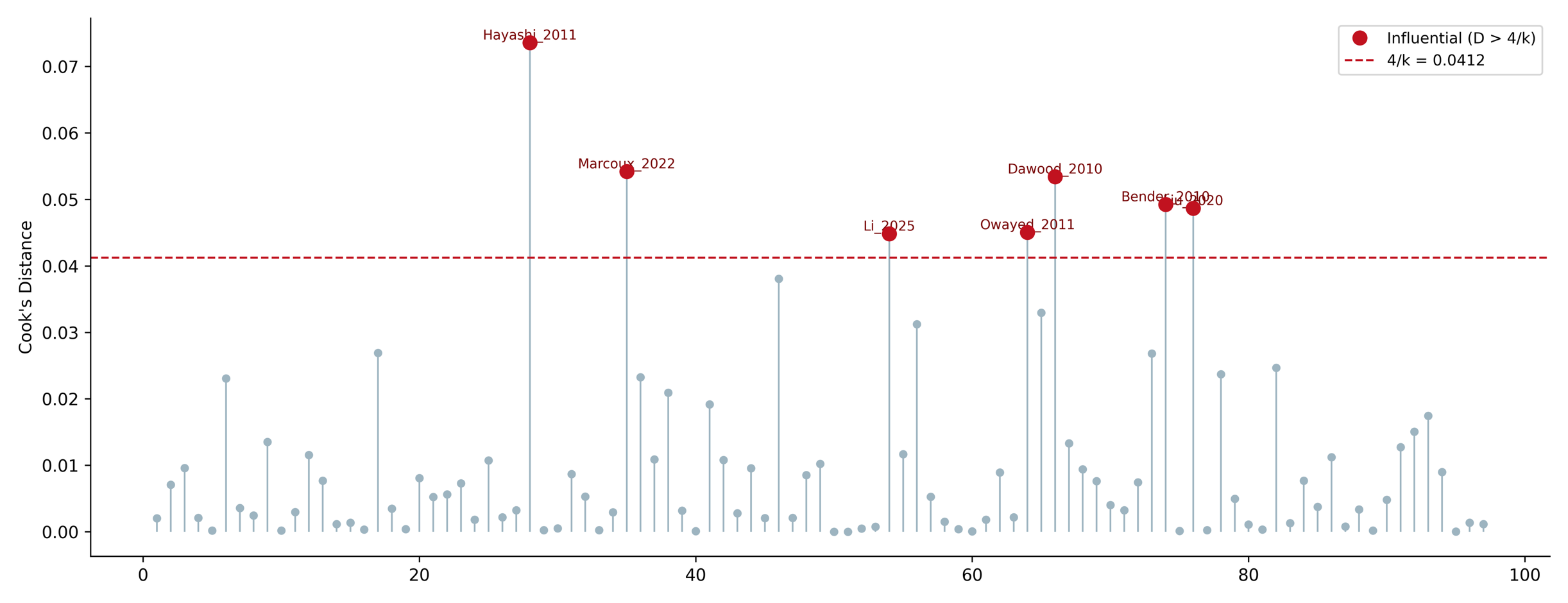

|  | **Pooled Prevalence (%)** | **95% CI** | **I^2^** |
| --- | --- | --- | --- |
| Full (n=97) | 16.7% | 13.9-20.0 | 99.0 |
| Reduced (n=90) | 17.9% | 15.3-20.8 | 98.3 |

**Supplementary Figure 1 - Influence diagnostics using Cook’s distance for included studies.**

Cook’s distance values are shown for all studies included in the meta-analysis (n = 97) to assess the influence of individual studies on the pooled prevalence estimate. The horizontal dashed line represents the predefined influence threshold (0.0412). Studies exceeding this threshold are labelled and considered potentially influential contributors to between-study heterogeneity and the overall pooled effect. The majority of studies fall below the threshold, indicating that the pooled estimate is not driven by a single disproportionately influential study. Seven studies were identified during the calculation of Cook’s Distance, these were then excluded for a “Leave One Out” analysis. When tested with a two-tailed Wald Z-test there was deemed to be no significant difference (0.5677) between the two groups.

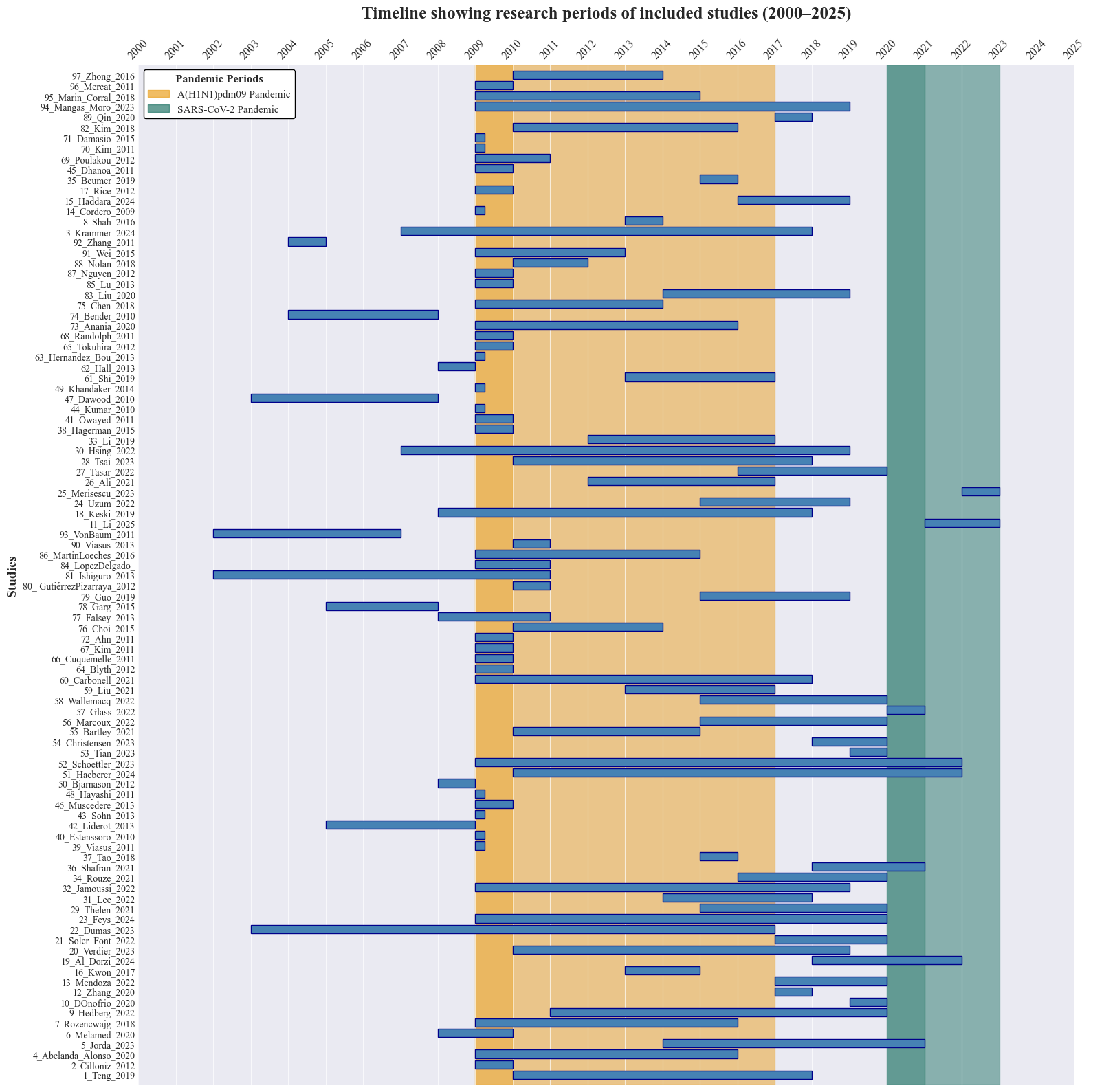

**Supplementary Figure 2 – Temporal distribution of research periods, with 2009 H1N1 and SARS-CoV-2 Pandemics highlighted.**

Darker yellow highlights the outbreak of the A/H1N1 2009 Pandemic strain, while the lighter yellow marks the use of the pandemic strain in influenza vaccines. The green highlight represents the COVID-19 Pandemic.
